# A data-driven Alzheimer’s disease progression simulator for retrospective validation and prospective Phase III power design

**DOI:** 10.64898/2026.05.03.26352317

**Authors:** Marco Lorenzi, Anna Custo, Giovanni B Frisoni, Valentina Garibotto

**Author notes:** joint first authors (Marco Lorenzi, Anna Custo).

## Abstract

Anti-amyloid immunotherapies have recently demonstrated the first significant slowing of cognitive decline in Alzheimer’s disease (AD), yet clinical benefit varies markedly across drugs and scales with the completeness of amyloid clearance. Pharmacokinetic/pharmacodynamic (PK/PD) models are currently the standard tool for trial simulation, but they typically operate on single biomarkers and rely on drug-concentration assumptions, leaving the multi-scale cascade from amyloid clearance through tau, neurodegeneration, and cognition largely unmodelled. No existing framework has been jointly validated against the quantitative outcomes of multiple real-world phase III trials, spanning clearance kinetics, multi-modal biomarker trajectories, and statistical power.

We present a trial simulation platform based on SimulAD, a disease progression model trained exclusively on longitudinal observational data from ADNI, with no access to trial-arm labels or drug-specific outcomes. SimulAD encodes intervention as piecewise amyloid clearance terms within a latent ordinary differential equation system that jointly governs amyloid, tau, structural MRI, and cognitive trajectories under the amyloid cascade hypothesis. We retrospectively simulated six landmark phase III anti-amyloid trials (TRAILBLAZER-ALZ2, CLARITY AD, EMERGE and ENGAGE, GRADUATE I and GRADUATE II) using a single trained model with trial-specific calibration limited to amyloid clearance kinetics.

SimulAD reproduced published mean centiloid reductions within 5% error across all six trials and generated CDR-SB distributions broadly consistent with reported placebo and treated-arm outcomes. In a retrospective power analysis, calibrated simulations separated the three positive from the three null trials, with EMERGE near the decision boundary and ENGAGE and both GRADUATE trials below it. Across trials, higher amyloid-clearance rates were associated with larger calibrated clinical effects and lower estimated sample sizes.

These results establish SimulAD as a valid disease-progression-centric trial simulator providing quantitative guidance on sample size planning and treatment kinetics optimisation that is grounded in the full multi-modal biomarker cascade of AD.

## 1. Introduction

Alzheimer’s disease (AD) affects an estimated 55 million people worldwide, a figure projected to nearly triple by 2050, imposing an escalating burden on patients, caregivers, and health systems globally (1). For more than two decades, the field witnessed an exceptional rate of clinical trial failures, with over 200 candidate disease-modifying therapies falling short in late-stage development (2). The recent phase III successes of lecanemab (CLARITY AD) and donanemab (TRAILBLAZER-ALZ2) have transformed this landscape: for the first time, amyloid immunotherapy demonstrably slowed cognitive and functional decline in early symptomatic AD (3; 4) Yet alongside these advances, unexplained discrepancies, such as the discordant outcomes in two identically designed trials of the same drug (aducanumab: EMERGE and ENGAGE), underscore a fundamental gap in our quantitative understanding of what determines trial success (5).

The positive trials have yielded two pivotal lessons about anti-amyloid intervention. First, cognitive benefit scales with the rate and completeness of amyloid clearance: TRAILBLAZER-ALZ2 achieved near-complete plaque removal (−88 centiloids at 18 months) and produced a 35% slowing of decline on the CDR-SB scale, while CLARITY’s more moderate clearance (−56 centiloids) yielded a 27% slowing at 18 months. Second, precise biomarker-guided patient selection targeting individuals with elevated amyloid pathology but not yet with advanced dementia appears critical to capturing the therapeutic window (6). Together, these findings validate the amyloid cascade hypothesis (7) and shift the central challenge of trial design from target identification to kinetics optimisation and population precision.

The kinetics of amyloid clearance are emerging as a primary determinant of cognitive outcomes. The EMERGE/ENGAGE discrepancy illustrates this clearly: despite identical protocols, real-world differences in baseline amyloid burden and dose delivery between populations likely explain why one trial met its primary endpoint and the other did not (5). More broadly, the causal chain from amyloid clearance to cognitive benefit involves a multi-scale cascade spanning tau propagation, structural neurodegeneration, and synaptic dysfunction, that unfolds over months to years and interacts non-linearly with a patient’s disease stage (8). Predicting how a specific clearance rate in a specific patient population will translate into a detectable clinical signal remains an unsolved quantitative problem.

Computational trial simulation offers a principled, scalable approach to address this challenge. Unlike real trials, in silico experiments allow systematic exploration of the full design space, varying amyloid clearance kinetics, patient baseline characteristics, sample sizes, and endpoint timing, at a fraction of the cost and duration (9). Pharmacokinetic/pharmacodynamic (PK/PD) modelling is currently the dominant paradigm for in silico trial simulation (10): by coupling drug-concentration kinetics to a pharmacodynamic biomarker response, PK/PD models can predict dose-response relationships and inform dosing schedules. However, PK/PD models of AD typically operate on a single pharmacodynamic endpoint (e.g. amyloid burden or a cognitive score) and do not capture the multi-scale cascade from amyloid clearance through tau propagation, structural neurodegeneration, and cognitive decline. Disease-progression models, by contrast, are designed to jointly track this full biomarker cascade, but are largely trained on natural history data and lack a principled mechanism for encoding intervention (11; 12; 13; 14). Critically, no existing approach, whether PK/PD or disease-progression-based, has been jointly validated against the quantitative outcomes of multiple phase III trials: amyloid clearance kinetics, multi-modal biomarker trajectories, and statistical power.

SimulAD is a variational autoencoder framework for multi-modal AD disease progression modelling, trained exclusively on longitudinal observational data from the Alzheimer’s Disease Neuroimaging Initiative (ADNI), with no access to trial-arm assignments or drug-specific outcome labels (15). By jointly learning the natural dynamics of amyloid PET, tau PET, structural MRI, and cognitive scores from time-series patient data, SimulAD can simulate counterfactual intervention scenarios without any interventional training data. This represents a potential critical advantage given the scarcity of labelled clinical trial datasets. Drug effects are introduced as additive clearance terms modifying the modelled dynamics, with parameters calibrated against published trial outcomes. Previous work demonstrated SimulAD’s ability to simulate natural AD progression and the cascading effects of amyloid intervention on downstream biomarkers, with independent validation on memory clinic cohorts (16).

In this work, we extend the SimulAD framework to retrospectively simulate six landmark anti-amyloid phase III trials: TRAILBLAZER-ALZ2 (donanemab), CLARITY AD (lecanemab), EMERGE and ENGAGE (aducanumab), and GRADUATE I and GRADUATE II (gantenerumab). A single computational model jointly accounts for amyloid, tau, structural, and cognitive biomarker trajectories across all six trials spanning a four-fold range in amyloid clearance rate. Kinetic calibration matched simulated clearance to published values within 5 percentage points across all six trials, and closely matched CDR-SB baseline and follow-up distributions for both placebo and treated arms. Power curves as a function of sample size reveal a tight coupling between amyloid clearance rate and the minimum sample size needed to detect a significant cognitive effect. SimulAD’s prospectively calibrated estimates correctly predicted the probability of statistical significance at the enrolled sample size, with all positive trials showing empirical power ≥ 65% (TRAILBLAZER 99%, CLARITY 86%, EMERGE 65%) and all negative trials showing empirical power ≤ 47% (ENGAGE 44%, GRADUATE II 47%, and GRADUATE I 24%). The EMERGE/ENGAGE discrepancy was also identified as a consequence of real-world differences in baseline amyloid burden, without requiring post-hoc adjustments.

Together, these results establish SimulAD as a validated, data-driven trial simulator offering quantitative guidance on sample size planning, population selection, and treatment kinetics optimisation grounded in the full multi-modal biomarker cascade of AD.

## 2. Results

### 2.1 Simulation of disease progression and drug-induced amyloid reduction

Our experimental approach is summarized in Figure 1, illustrating the use of SimulAD to simulate causal intervention based on target intervention scenarios. Starting from a training cohort composed by longitudinal multi-modal observations of amyloid, tau, structural brain imaging biomarkers and cognitive scores (panel A), SimulAD integrates this information to describe the long-term progression of Alzheimer’s disease, along with the dynamics of interactions between modalities across the disease time course (panel B). We simulated intervention in SimulAD by modifying the dynamics of amyloid accumulation according to prior clearance kinetics reported in clinical trials (panel C), to obtain hypothetical disease progression time courses corresponding to the prescribed intervention scenario (panel D). Finally, we estimated the associated downstream treatment efficacy by sampling the associated trajectories of clinical scores in simulated controls and placebo arms, and by quantifying the calibrated effect size and statistical power (panel E).

**Figure 1.**
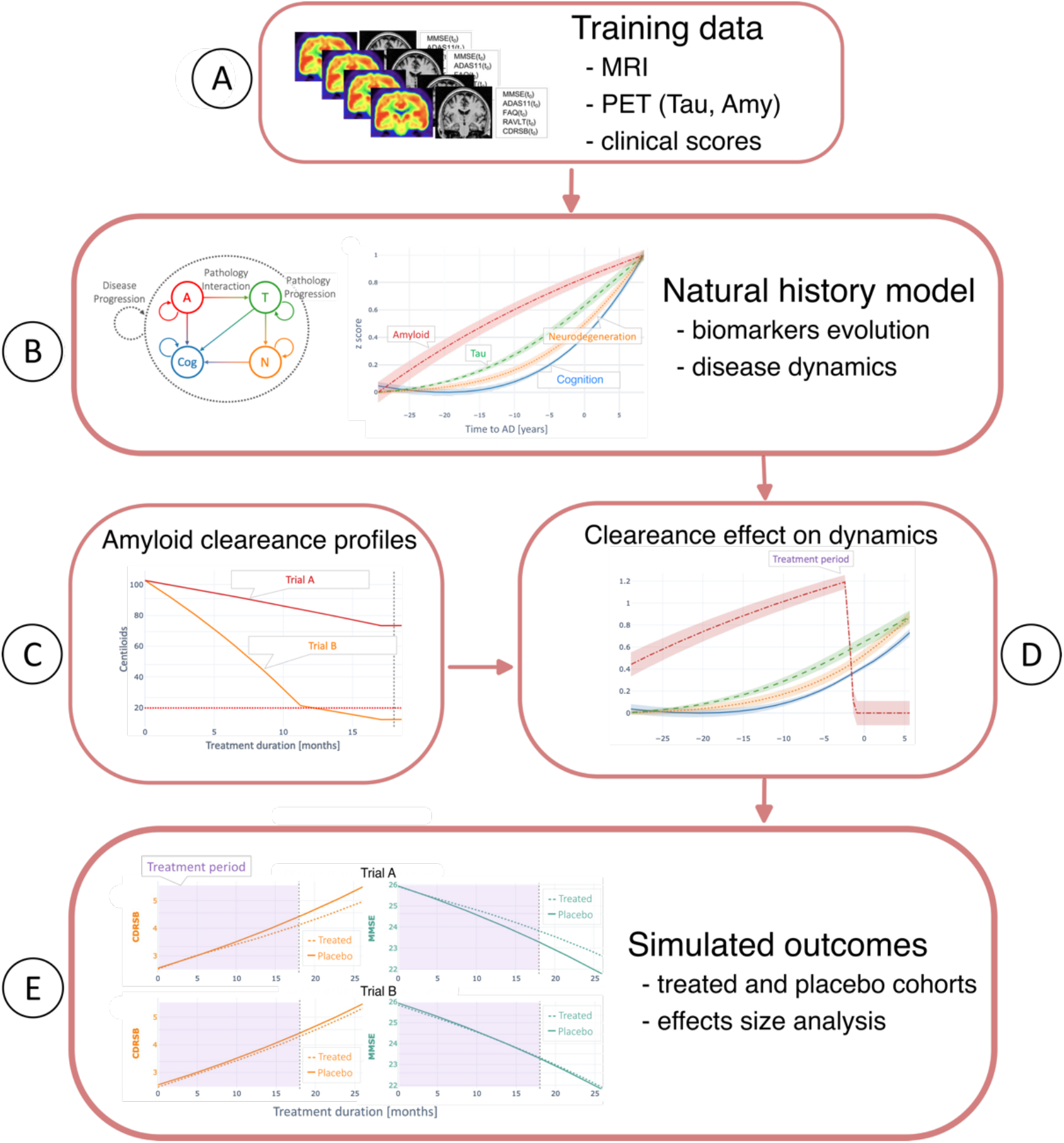
Our experimental approach. A. Training cohort composed by longitudinal multi-modal observations of amyloid, tau, structural brain imaging biomarkers and cognitive scores. B) Long-term progression of Alzheimer’s disease estimated by SimulAD, along with the dynamics of interactions between modalities across the disease time course. C) Prior clearance kinetics reported in clinical trials. D) We simulated intervention in SimulAD by modifying the dynamics of amyloid accumulation according to prior kinetics. E) Estimated treatment on the associated trajectories of clinical scores in simulated controls and placebo arms.

### 2.2 Calibration of amyloid clearance kinetics

Drug-induced amyloid clearance was modelled by a piecewise-constant rate function *γ*(t) applied additively to the AV45 latent dimension (see Methods, Eq. 2). The piecewise parameterisation captures the clinically observed phenomenon whereby clearance rates are highest early in treatment and decrease progressively as the burden declines. Each trial was assigned a trial-specific phase structure based on its published dosing and titration schedule: the time points at which the active clearance rate transitions to a lower value were aligned with titration milestones or the anticipated onset of plateau kinetics inferred from reported centiloid trajectories.

The *γ* parameters were calibrated iteratively against published centiloid values at the primary endpoint (see Methods). At each iteration, the ratio k = *γ*_n+1_ / *γ*_n_ was computed, where percentage centiloid clearance is defined as %CL = (CL_0_ − CL_FU) / CL_0_ × 100. *γ* values were scaled by k. We adopted fractional rather than absolute clearance as the calibration target to avoid confounds between the simulated and real populations arising because SimulAD draws subjects from the ADNI cohort rather than the trial-specific enrolment population. Rapid convergence (*Δ*%CL < 2 pp) was achieved in two to five iterations per trial.

Table 1 summarises the calibrated *γ* parameters and the resulting centiloid clearance at the primary endpoint for each of the six trials. The calibrated initial clearance rates *γ*_0_ span more than four-fold, from *γ*_0_ = 0.654 for donanemab (TRAILBLAZER-ALZ2), to *γ*_0_ = 0.151 for gantenerumab (GRADUATE I/II). Lecanemab (CLARITY AD) required a three-phase model, with phase transitions at months 6, 12, and 18 and rates *γ* = {0.468, 0.200, 0.065}, to reproduce the characteristic deceleration of clearance observed in the published centiloid time-course, in which clearance accelerates sharply through month 6 but slows appreciably thereafter as accessible amyloid pools are progressively depleted. Aducanumab (EMERGE, ENGAGE) showed intermediate initial kinetics (*γ*_0_ = 0.249 and 0.189, respectively), with a sustained secondary phase at a comparable rate extending to month 18, consistent with its progressive removal of fibrillar deposits. Gantenerumab’s substantially lower *γ*_0_ reflects both its slower dose-titration schedule and the longer 27-month treatment period required to achieve comparable absolute centiloid reductions.

**Table 1.**
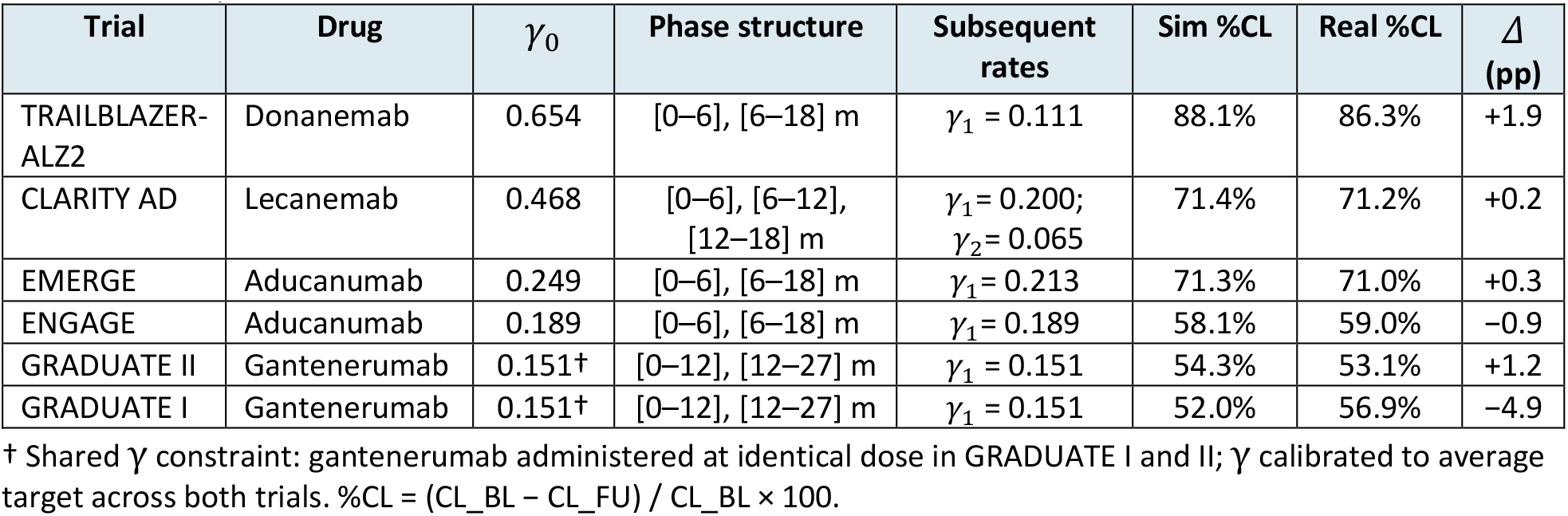
Calibrated piecewise clearance rate parameters *γ*(t) and calibration accuracy. *γ*_0_ = initial clearance rate; Sim %CL = simulated fractional centiloid clearance at primary endpoint; Real %CL = published value; Δ (pp) = residual error. † Shared *γ* constraint (identical drug and dose in GRADUATE I/II).

| Trial | Drug | $\gamma_0$ | Phase structure | Subsequent rates | Sim %CL | Real %CL | $\Delta$ (pp) |
| --- | --- | --- | --- | --- | --- | --- | --- |
| TRAILBLAZER-ALZ2 | Donanemab | 0.654 | [0–6], [6–18] m | $\gamma_1 = 0.111$ | 88.1% | 86.3% | +1.9 |
| CLARITY AD | Lecanemab | 0.468 | [0–6], [6–12], [12–18] m | $\gamma_1 = 0.200$; $\gamma_2 = 0.065$ | 71.4% | 71.2% | +0.2 |
| EMERGE | Aducanumab | 0.249 | [0–6], [6–18] m | $\gamma_1 = 0.213$ | 71.3% | 71.0% | +0.3 |
| ENGAGE | Aducanumab | 0.189 | [0–6], [6–18] m | $\gamma_1 = 0.189$ | 58.1% | 59.0% | –0.9 |
| GRADUATE II | Gantenerumab | 0.151† | [0–12], [12–27] m | $\gamma_1 = 0.151$ | 54.3% | 53.1% | +1.2 |
| GRADUATE I | Gantenerumab | 0.151† | [0–12], [12–27] m | $\gamma_1 = 0.151$ | 52.0% | 56.9% | –4.9 |

† Shared $\gamma$ constraint: gantenerumab administered at identical dose in GRADUATE I and II; $\gamma$ calibrated to average target across both trials. $\%CL = (CL_{BL} - CL_{FU}) / CL_{BL} \times 100$ .

The piecewise *γ* profiles and resulting simulated centiloid trajectories are illustrated in Figure 2. The ordering of *γ* values recapitulates the established clinical hierarchy of amyloid-lowering efficacy: donanemab ≫ lecanemab ≈ aducanumab (EMERGE) > aducanumab (ENGAGE) > gantenerumab.

**Figure 2.**
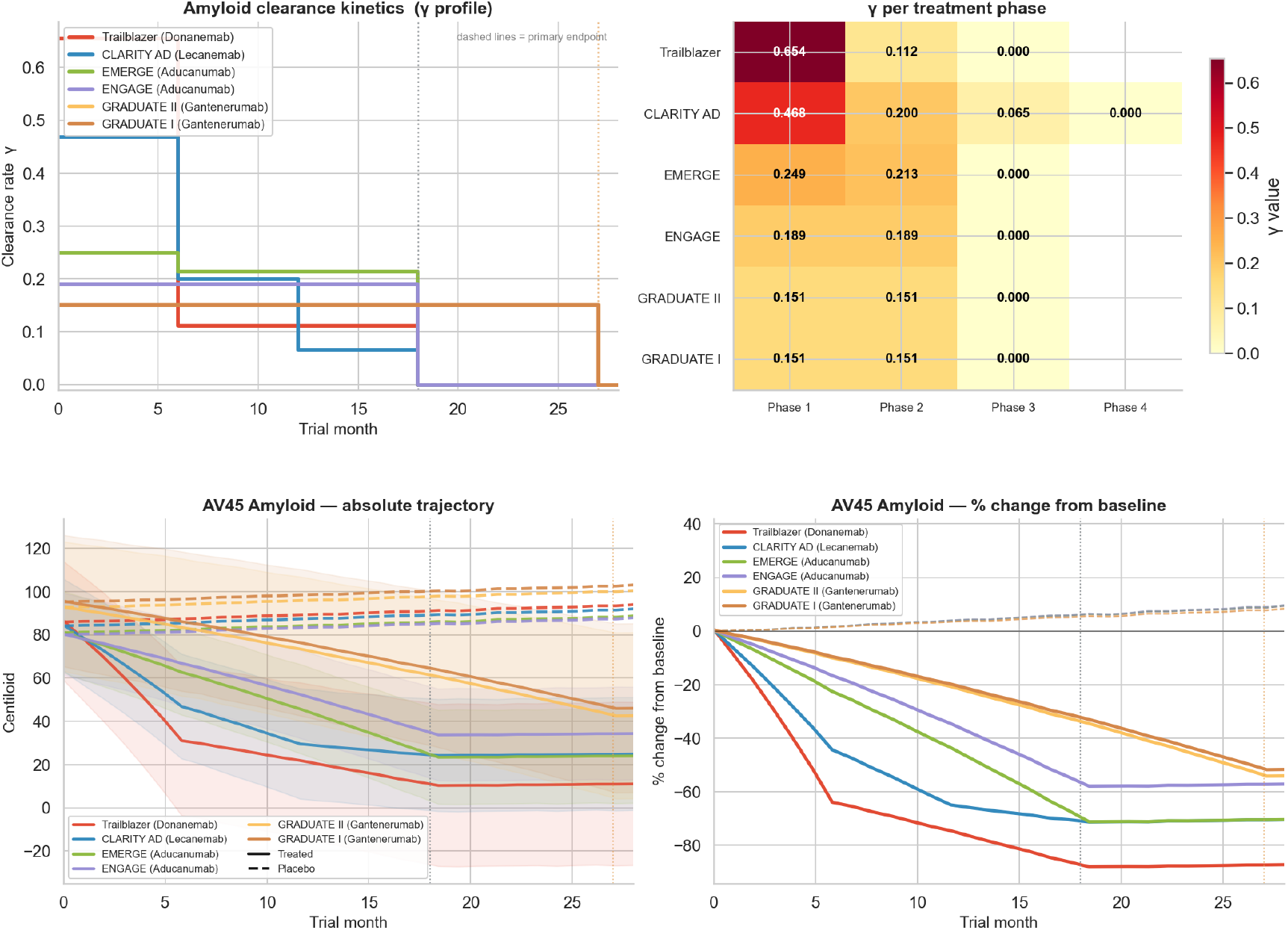
Calibrated amyloid clearance kinetics across six anti-amyloid immunotherapy trials. (top) Piecewise-constant clearance rate *γ* profiles as a function of time from treatment onset. Vertical dashed lines mark phase transitions. The ordering of *γ* values recapitulates the clinical hierarchy of amyloid-lowering strength: donanemab (*γ* = 0.654) > lecanemab (*γ* = 0.468) > aducanumab EMERGE (*γ* = 0.249) > aducanumab ENGAGE (*γ* = 0.189) > gantenerumab I/II (*γ* = 0.151). (bottom) Simulated centiloid trajectories (mean ± SD, 100 repetitions × 150 subjects/arm) for treated (filled symbols) and placebo (open symbols) arms. Shaded bands: ±1 SD across repetitions.

To validate the calibration, simulated absolute centiloid values at baseline and primary endpoint were compared head-to-head with published values for both arms (Figure 3). At the primary endpoint, the simulated treated-arm centiloid agreed with published figures within 5 CL for five of six trials: TRAILBLAZER-ALZ2 (sim 10.3 vs real 14.0 CL; Δ = −3.7 CL), CLARITY AD (24.2 vs 22.4 CL; Δ = +1.8 CL), EMERGE (23.3 vs 24.6 CL; Δ = −1.3 CL), ENGAGE (33.7 vs 37.3 CL; Δ = −3.6 CL), and GRADUATE II (42.5 vs 44.9 CL; Δ = −2.4 CL). In fractional terms of amyloid clearance %CL, calibration accuracy was within 2 percentage points for these five trials (gaps: +1.9, +0.2, +0.3, −0.9, and +1.2 pp, respectively).

**Figure 3.**
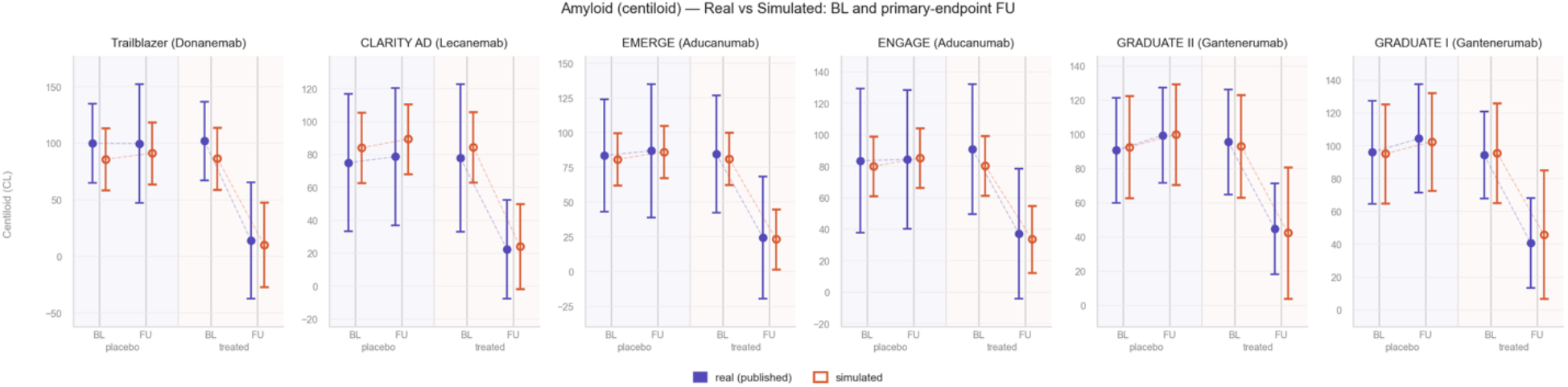
Head-to-head comparison of simulated and published centiloid values at baseline and primary-endpoint follow-up. For each trial, mean ± SD centiloid is shown for treated and placebo arms at baseline (BL) and primary-endpoint follow-up (FU). Orange whiskers: SimulAD simulation (100 repetitions × 150 subjects/arm). Blue whiskers: published trial values. Placebo arm values reflect natural ADNI amyloid accumulation trajectory and SimulAD does not model drug effects on the placebo arm.

The largest residual discrepancy was observed for GRADUATE I (simulated follow-up (FU) centiloid 45.9 vs published 40.7 CL; fractional clearance 52.0% vs published 56.9%; gap −4.9 pp). This reflects a deliberate pharmacological constraint: because gantenerumab was administered at an identical dose in both GRADUATE I and GRADUATE II, a single *γ* value was assigned to both trials and calibrated to their average clearance target. This constraint is physiologically motivated and results in an asymmetric residual error that mildly underestimates GRADUATE I clearance (−4.9 pp) while slightly overestimating GRADUATE II (+1.2 pp). Decoupling *γ* between the two trials would reduce the GRADUATE I error but would introduce a degree of freedom inconsistent with the shared pharmacology.

Temporal calibration fidelity was further assessed against the intermediate centiloid time-points reported at months 3, 6, 12, and 18 in the CLARITY AD primary publication (3). The simulated trajectory reproduced these values within 6 CL at all four assessments (Δm3 = +2.3 CL, Δm6 = −2.7 CL, Δm12 = −5.0 CL, Δm18 = −5.3 CL), corresponding to simulated fractional clearance of 21.0%, 44.6%, 65.1%, and 71.4% against published values of approximately 25.7%, 44.9%, 64.2%, and 70.6%. The three-phase model thus captures both the rapid initial phase (months 0–6, *γ* = 0.468) and the following deceleration in the second year of treatment (months 12–18, *γ* = 0.065) that characterise lecanemab’s clearance mechanism.

### 2.3 Simulated CDR-SB trajectories and statistical power analysis

#### Head-to-head CDR-SB comparison

SimulAD simulations reproduce the qualitative pattern of CDR-SB progression with increasing scores in the placebo arm and attenuated progression in the treated arm. Figure 4 shows the reconstructed absolute CDR-SB at the primary-endpoint follow-up for treated and placebo arms under the adopted fully prospective calibration (see Methods Section 4.5). In this scenario, the model computed the baseline CDR-SB scores based on the calibrated simulation baseline placebo targets (Section 4.3.1). Placebo and treated follow-up predictions were computed from prospectively calibrated SimulAD predictions according to the procedure detailed in Section 4.5.4. Importantly, this calibration strategy is implemented without requiring target endpoint data from the target trial. The simulations produced consistent treated-arm predictions for the six trials coherent with the real-world values reported in the respective target trials. In Supplementary Section S4 we show the calibration parameters and related simulations’ results across different calibration strategies. In spite of the less restrictive assumptions on prior trial knowledge, all calibrations lead to similar predictions’ accuracy for CDRB-SB across trials.

**Figure 4.**
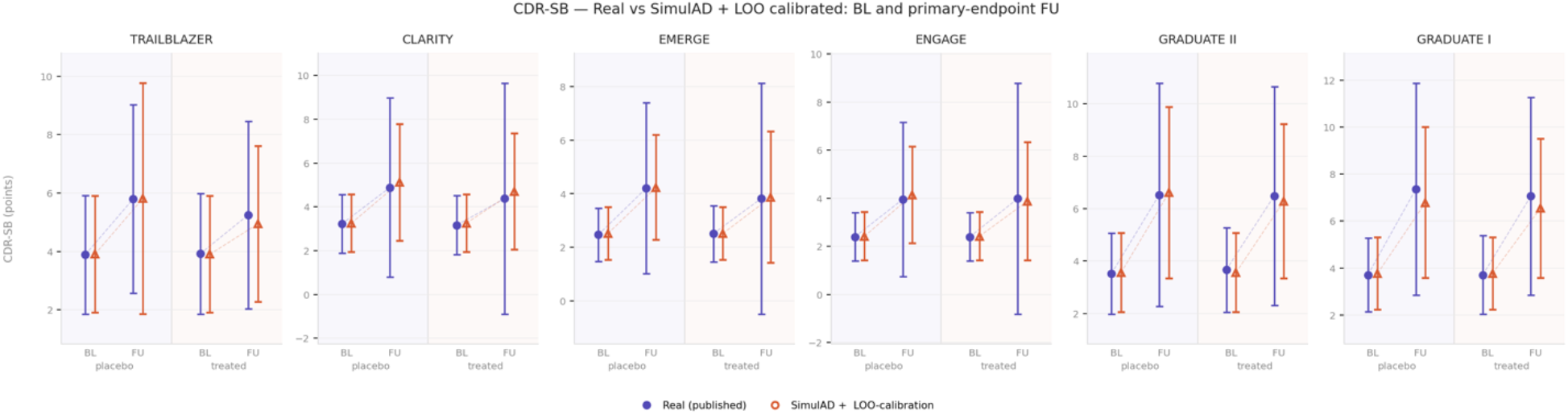
Calibrated SimulAD CDR-SB predictions versus published trial values at baseline (BL) and primary-endpoint follow-up (FU). For each trial, mean CDR-SB ± 1 SD is shown for placebo (left) and treated (right) arms. Published values (blue circles) are compared with SimulAD prospectively calibrated predictions (orange triangles) (Section 4.5.4).

#### Calibrated power and sample size

Figure 5 shows calibrated power curves and effect etimates, while Table 2 summarises N80 and power at trial N estimated. The calibrated framework correctly identifies the three positive trials: TRAILBLAZER and CLARITY achieve ≥80% power at their actual enrolled N, while EMERGE is borderline (65%), in line with the reported modest significance in the original trial (p = 0.01, (5)). On the contrary, ENGAGE, GRADUATE II, and GRADUATE I are estimated at <50% power, consistent with their published null results (5; 17). Similar results are obtained under the alternative calibration schemes, while raw simulations overestimate power for GRADUATE II (53% vs oracle 18%) and underestimate for EMERGE (55% vs oracle 84%) (Supplementary Section S5).

**Table 2.** Calibrated N80 estimates for 80% power (*α* = 0.05, two-sided). Oracle: N80 from published real effect and SD. Power@N_actual: empirical power at the enrolled sample size, interpolated from the simulated N grid.

| Trial | Oracle $N_{80}$ | SimulAD $N_{80}$<br>(LOO calibrated) | Power@N<sub>actual</sub><br>(LOO calibrated) |
| --- | --- | --- | --- |
| TRAILBLAZER | 333 | 148 | 99% |
| CLARITY | 600 | 668 | 86% |
| EMERGE | 500 | 722 | 65% |
| ENGAGE | >4000 | 1349 | 44% |
| GRADUATE II | 3658 | 1324 | 47% |
| GRADUATE I | 1375 | 2269 | 24% |

**Figure 5.**
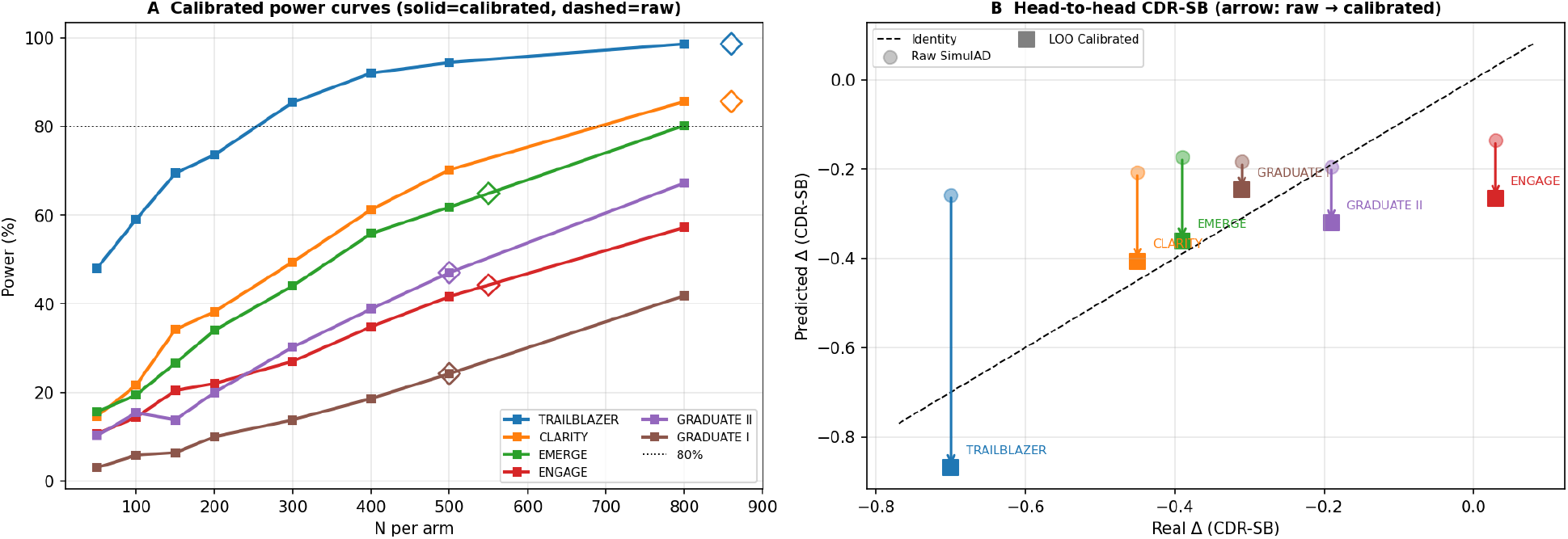
Panel A. Calibrated empirical power curves for six anti-amyloid phase III trials, shown on a single plot. Each line gives the fraction of 500 simulated trials achieving significance (p < 0.05, two-sided) after calibration as a function of N per arm. Diamonds mark the actual enrolled sample size for each trial; the horizontal dotted line marks the 80% power threshold. Panel B. Head-to-head comparison of SimulAD treatment effect estimates against published trial values (CDR-SB points). Each trial is shown as a raw SimulAD prediction (open circle, mean over 500 repetitions at N = 800 per arm) and a LOO-calibrated prediction (filled square); arrows indicate the direction and magnitude of the calibration correction. The dashed line is the identity. Points on or near the identity line indicate accurate prediction; the calibration moves all trials closer to the identity except ENGAGE, where the LOO multiplier overcorrects.

### 2.4 Comparison with simpler approaches

Table 3 compare SimulAD’s prospective *N*_80_ estimates with the three reference approaches (Methods Section 4.6).

**Table 3.** Benchmark N80 comparison. Oracle: N80 from published real data. B1(sim): raw SimulAD effect + sim SD. B1(LOO): raw SimulAD effect + LOO estimated SD. B2: Ridge regression. B3: gradient-boosted model. >4000 indicates N80 exceeds 4000 (effectively unpowered or wrong direction).

| Trial | Oracle | SimulAD<br>prosp. | B1<br>(sim SD) | B1<br>(LOO SD) | B2<br>(Ridge) | B3<br>(GBM) |
| --- | --- | --- | --- | --- | --- | --- |
| TRAILBLAZER | 333 | 148 | 553 | 1675 | 1311 | 20 |
| CLARITY | 600 | 668 | 802 | 2561 | 3112 | 34 |
| EMERGE | 500 | 722 | 938 | 3152 | 2278 | 484 |
| ENGAGE | >4000 | 1349 | 1526 | >4000 | 2855 | 65 |
| GRADUATE II | 3657 | 1324 | 1093 | 3567 | >4000 | 41 |
| GRADUATE I | 1374 | 2269 | 1260 | 4121 | 3872 | 108 |

#### B1 baselines

B1 (sim SD) systematically overestimates N80 for the powered studies TRAILBLAZER (553 vs 333), CLARITY (802 vs 600) and EMERGE (938 vs 500). Substituting the LOO SD (B1-real) dramatically worsens accuracy across all trials (N80 range 1675–>4000 per arm), confirming that variance calibration alone, without effect-size calibration, amplifies rather than reduces estimation error.

#### B2 and B3 baselines

B2 (Ridge) predicts a positive treatment direction for all six trials, yielding N80 >1300 throughout, expected given the absence of an explicit intervention variable in its training data. B3 (GBM) shows high variance, occasionally close to oracle for individual trials (EMERGE: 484 vs 500) but without systematic accuracy and overconfident for the positive trials (TRAILBLAZER: 20, CLARITY: 34).

Overall, SimulAD prospective calibration achieves the closest agreement with oracle N80 across all six trials. The advantage derives from coupling mechanistic amyloid-clearance kinetics (k∼*Δ*CL calibration) with a prospective variance estimate: neither component alone is sufficient, as the B1-real result demonstrates.

### 2.5 Multimodal cascade analysis

The latent ODE coupling structure learned from ADNI shows that the direct W[AV45→MRI] weight is exactly zero, while MRI is affected only via the two-step indirect cascade AV45→TAU→MRI (W[AV45→TAU] = −3.7×10^−4^, W[TAU→MRI] = +4.3×10^−2^; composite coupling −1.6×10^−5^). These interaction dynamics learned by SimulAD determine the simulated biomarkers relationship. Individual amyloid decrease correlated more strongly with entorhinal tau reduction than with hippocampal volume change. In the treated arm (pooled across trials after within-trial z-scoring), Pearson r = +0.46 (Spearman ρ = +0.44) for ΔAV45 vs ΔTAU, and r = −0.20 (ρ = −0.19) for ΔAV45 vs ΔVol. In the placebo arm, natural-history covariance was substantially stronger: r = +0.71 (ρ = +0.70) for tau and r = −0.70 (ρ = −0.68) for volume. Per-trial correlation analysis is provided in Supplementary Section S7. Overall, the treatment-arm attenuation relative to placebo reflects treatment-driven amyloid variance captured by SimulAD that is partially decorrelated from the natural-history biomarker trajectories.

## 3. Conclusions and Discussion

The central finding of this work is that SimulAD faithfully reproduces the quantitative outcomes of six landmark anti-amyloid phase III trials, correctly classifying the significance of all six after LOO-calibrated correction of both effect size and pooled standard deviation.

A key quantitative result is the coupling between amyloid clearance rate and required sample size. Prospective *N*_80_ estimates increase monotonically as *Δ*CL decreases from 88% (TRAILBLAZER, *N*_80_ = 148) to 60% (EMERGE, 722) to 46% (GRADUATE II, 1324). This scaling follows directly from the k∼*Δ*CL/trial_duration calibration. For GRADUATE II, 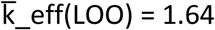 falls below k_sd(LOO) = 1.78, giving k_ratio = 0.92 < 1, so the calibrated Cohen’s d is smaller than the raw simulated Cohen’s d, correctly predicting higher N_80_ and lower power than the raw simulation suggests. This suggests a practical threshold: *Δ*CL ≈ 55–60% separates under-powered from adequately-powered 27-month trials at current effect-size levels. Taken together, our results show that high-clearance drugs trials are potentially over-designed. Had SimulAD been available at the design stage, it would have enabled leaner, faster trials. On the contrary, moderate-clearance drugs trials are critically under-designed at standard Phase III sizes. SimulAD’s most valuable contribution would have been flagging that N_80_ at this clearance level is in the thousands, converting a go/no-go decision before committing to Phase III.

It is worth adding some clarifying remarks on the *N*_80_ estimates. First, TRAILBLAZER-ALZ2 enrolled approximately 860 participants per arm, yet the calibrated model estimates *N*_80_ = 148: the trial was therefore approximately 3 times overpowered for its primary CDR-SB endpoint. This deliberate over-enrolment provided statistical margin for secondary biomarker sub-studies, safety endpoints, and the pre-specified disease-modification sub-group analysis, and is consistent with the pragmatics of Phase III design where statistical risk must be minimised for a registrational programme.

Second, the ENGAGE oracle *N*_80_ is effectively undefined: the published placebo–treated CDR-SB difference at 18 months was approximately +0.03, which projects to a required sample size largely exceeding 10 000 per arm at 80% power. Therefore, SimulAD’s calibrated N_80_ for ENGAGE (≈ 1,300 per arm, fully prospective) reflects the enrolment that would be required if the aducanumab mechanism had produced the model-predicted effect, and not the sample size needed to replicate the real null outcome. For this reason, because ENGAGE’s real effect is near zero and of uncertain sign, it was excluded from the calibration set and its 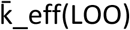 is estimated out-of-sample from the five-trial regression.

Third, GRADUATE I and GRADUATE II are simulated with identical gantenerumab kinetics (Section 2.2), but their published amyloid clearances differ, reflecting a trial-level compromise in the simulation. The k ∼ ΔCL/trial_duration LOO regression uses the published clearance values, yielding different 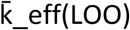 predictions (1.64 vs 1.35) and calibrated N_80_ estimates (1,324 vs ∼2,269 per arm). This difference in calibrated N_80_ therefore captures the real-world difference in drug performance between the two trials, not a calibration artefact.

Finally, the 50% threshold adopted in this work is a classification device, not a sample-size design standard. This threshold quantifies whether a trial, under the calibrated simulator, is likely to achieve nominal significance at its enrolled sample size. This is distinct from *N*_80_, which remains the appropriate prospective design criterion. The proximity of EMERGE (64.9%) and GRADUATE II (47.0%) also indicates that the classification margin is modest for some trials.

The tight linear relationship between 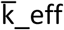 and *Δ*CL/t (*R*^2^ = 0.85, Supplementary Section S6) has a natural interpretation. SimulAD’s ODE dynamics are learned from ADNI natural-history trajectories, which sample the amyloid–tau–cognition cascade in a regime of slow, moderate amyloid accumulation (*Δ*CL ≈ 0 in untreated subjects over 18–27 months). The inter-modality coupling coefficients are therefore estimated from marginal dynamics near the natural-history attractor. When a drug displaces the system far from this attractor, as for example with donanemab, the ODE integrates through a regime of cascade dynamics unseen during training. In this out-of-distribution regime the learned coupling coefficients systematically underestimate the downstream cognitive amplification. 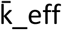 thus acts as an out-of-distribution correction factor: the further the intervention pushes the system from the natural-history attractor, the larger the correction required. A complementary source of model error captured by 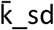, is a systematic underestimation of CDR-SB inter-subject variability. ADNI participants are selected for protocol compliance, relatively homogeneous disease staging, and frequent clinical contact, producing lower between-subject dispersion in CDR-SB trajectories than the broader, more heterogeneous populations enrolled in regulatory trials. The scalar 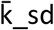 correction compensates for this gap but does not identify its root causes. Overall, both 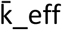 and 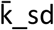 should be treated as an empirical prior subject to revision as new Phase 3 trial data become available.

Beyond sample size planning, SimulAD can directly inform patient population selection. By varying the clinical subgroup from which the simulated population is drawn, for example by restricting to MCI-to-AD converters versus a broader early MCI population, the framework predicts how baseline amyloid burden and disease stage affect both amyloid clearance dynamics and the detectable CDR-SB treatment effect. The results suggest that populations with higher baseline amyloid burden achieve faster effective clearance under the same drug, translating into larger treatment effects at smaller sample sizes. These population-level insights can support biomarker-guided enrichment strategies, such as minimum centiloid thresholds for inclusion or maximum tau-PET thresholds to exclude advanced neurodegeneration, without requiring any additional trial-specific training data. In practical terms, the calibrated power curves in Figure 5 quantify the efficiency gains achievable by targeting interventions with higher amyloid clearance rates. These results define a concrete prospective workflow for Phase 3 design from Phase 2 data.

Concerning the mechanistic interpretation of the simulations, we note that SimulAD predictions are broadly consistent with the available clinical evidence from anti-amyloid trials. For tau pathology, a pre-specified substudy of CLARITY AD showed slowed brain tau plaques accumulation relative to placebo beyond 18 months (18). The direction of the SimulAD prediction (small but non-zero tau slowing, scaling with amyloid clearance) is qualitatively aligned with this finding, though the magnitude comparison is confounded by the population variability and trials modalities. For structural MRI, clinical evidence from anti-amyloid trials has not uniformly demonstrated hippocampal volume preservation. This mixed picture is consistent with SimulAD’s prediction of negligible hippocampal volume preservation. Taken together, the multimodal cascade analysis reveals that SimulAD’s primary mechanistic limitation is not in the tau–cognition or MRI–cognition coupling (W[TAU→CLINIC] and W[MRI→CLINIC] are of appropriate sign and magnitude), but rather in the amyloid–tau cascade amplification. The calibration factor can therefore be understood as correcting not only for out-of-distribution amyloid dynamics, but also for the missing downstream amplification through the tau–cognition pathway that the trained ODE underestimates when amyloid is removed far beyond the natural-history range.

Several limitations of this study must be acknowledged. First, the LOO calibration factors 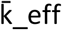 and 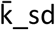 are scalar multipliers estimated from at most five trials. Their generalisation to future trials with substantially different patient populations, treatment protocols, or clearance profiles is untested. The regression should therefore be regarded as a calibration curve valid over the current *Δ*CL/t range, not as a mechanistic law, while extrapolation to substantially higher or lower clearance profiles carries proportionally larger uncertainty. Second, although 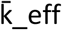 corrects for systematic LMM underestimation, the underlying source of the gap between ADNI-derived dynamics and trial populations remains uncharacterised. An additional statistical caveat concerns the k ∽ *Δ*CL/t regression itself (*R*^2^ = 0.85, Supplementary Figure S6), since this correlation is estimated from only five calibration data points. Sensitivity analysis confirms that the regression slope is robust to the exclusion of TRAILBLAZER (slope change from 0.48 to 0.68, Supplementary Section S4). Nevertheless, the increase of prediction-interval width after excluding TRAILBLAZER from the calibration (mean PI 2.15 → 3.18) underscores that the apparent precision of k predictions in the five-trial fit is partly driven by this single leverage point, and that k uncertainty for trials outside the observed ΔCL/t range may be larger. We also note that the amyloid clearance model is phenomenological: *γ*(t) is a piecewise-constant rate with no pharmacokinetic/pharmacodynamic (PK/PD) component, preventing direct mapping to drug dose, titration schedules, or ARIA risk, and precluding simulation of dose-response curves. Moreover, the centiloid calibration was performed using AV45 (florbetapir) PET data from ADNI, whereas the five trials used different tracers (florbetaben, PiB, florbetapir) with distinct SUVR-to-centiloid conversion factors, introducing additional calibration uncertainty. SimulAD does not model treatment-related side effects such as amyloid-related imaging abnormalities (ARIA), which are a critical safety consideration for anti-amyloid therapies and may influence discontinuation rates and effective exposure. Finally, the model is trained on the ADNI cohort, which is predominantly white, US-based, and research-engaged, and is therefore subject to potential selection bias toward this specific population. Although previous studies have demonstrated that the disease progression dynamics learned from ADNI generalise to independent memory clinic cohorts (16), broader validation on more ethnically and geographically diverse populations is essential. Real-world clinical populations differ from ADNI in comorbidity burden, education level, and amyloid measurement protocols, all of which may affect model calibration and the generalisability of its predictions.

To conclude, the demonstrated ability of SimulAD to calibrate amyloid clearance kinetics within 5% error, reproduce CDR-SB trajectories, correctly classify all six trial outcomes, and partially recover the EMERGE/ENGAGE discrepancy as a consequence of calibrated dynamics establishes a new quantitative benchmark for computational Alzheimer’s disease trial simulation. Together with the fully prospective *N*_80_ estimates and benchmark comparisons, these results position SimulAD as a practical complement to PK/PD modelling for Phase III design.

## 4. Methods

### 4.1 Data

Data used in the preparation of this article were obtained from the Alzheimer’s Disease Neuroimaging Initiative (ADNI) database (adni.loni.usc.edu) (19). The ADNI was launched in 2003 as a public-private partnership, led by Principal Investigator Michael W. Weiner, MD. The original goal of ADNI was to test whether serial magnetic resonance imaging (MRI), positron emission tomography (PET), other biological markers, and clinical and neuropsychological assessment can be combined to measure the progression of mild cognitive impairment (MCI) and early Alzheimer’s disease (AD). The current goals include validating biomarkers for clinical trials, improving the generalizability of ADNI data by increasing diversity in the participant cohort, and to provide data concerning the diagnosis and progression of Alzheimer’s disease to the scientific community. For up-to-date information, see adni.loni.usc.edu.

Specifically, we used the PET imaging biomarkers from UC Berkeley’s analysis and the MRI-based biomarkers, clinical scores, and diagnostics data available in ADNI MERGE (Supplementary Table S1). We included individuals with at least one positive CSF amyloid measure (CSF Abeta <182 pg/ml) and all imaging biomarkers (amyloid PET, tau PET, MRI) and clinical tests acquired during the study. As a result, we trained SimulAD on a cohort comprising 1256 data points from 351 individuals (Centiloid mean (std): 58.35 (43.40), Tau Fusiform SUVr mean (std): 1.35 (0.31), Intracranial volume (ICV) normalized brain volume mean (std): 0.71 (0.06), CDR-SB mean (std): 1.30 (2.11), MMSE mean (std): 27.6 (3.1), ADAS-13 mean (std): 14.30 (10.1)) (Supplementary Section S1).

### 4.2 Model Specification

SimulAD is a variational autoencoder (VAE) (20) coupled with a neural ordinary differential equation (ODE) (21) trained to reconstruct and predict longitudinal multi-modal biomarker trajectories. Four data modalities are considered: clinical scores (CLINIC), structural MRI (MRI), tau PET (TAU), and amyloid PET (AV45), each represented by several biomarker features (Section 4.1).

#### 4.2.1 Encoding

Each modality m is encoded by a dedicated linear layer that maps the observed baseline biomarkers *x*_m_ to a one-dimensional Gaussian posterior 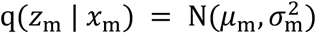, parameterised by mean *μ*_m_ and log-variance log(σ_m_). Encoder weights are initialised near zero (std = 0.001) and the log-variance bias is fixed to −20, encouraging near-deterministic initial embeddings. The four latent coordinates are concatenated into z ∈ ℝ^4^.

#### 4.2.2 Decoding

Each modality is reconstructed by a dedicated linear decoder 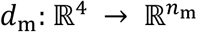. Decoder weights are initialised with 90% sparsity. The reconstruction likelihood uses a Gaussian noise model with a per-biomarker learnable variance 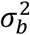, jointly optimised with all model parameters.

#### 4.2.3 Latent ODE Dynamics

Following (15), the temporal evolution of z is governed by a neural ODE (21):

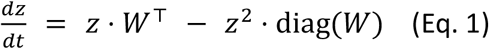

where *W* ∈ ℝ^4×4^ is a learnable weight matrix and diag(*W*) is its diagonal. The linear term *z* · *W*^T^ captures inter-modality coupling; the quadratic term provides self-saturation that bounds z-score trajectories. Integration uses a fixed-step midpoint solver (https://github.com/rtqichen/torchdiffeq).

Biologically motivated pathway constraints are imposed by masking off-diagonal entries of *W*. Only the following directed interactions are permitted: AV45→AV45, AV45→TAU, TAU→TAU, TAU→MRI, MRI→MRI, AV45→CLINIC, TAU→CLINIC, MRI→CLINIC, reflecting the hypothesized canonical amyloid cascade hypothesis (Figure 7). A quantitative representation of the dynamics learned by SimulAD on the ADNI cohort is provided in Supplementary Section S2.

**Figure 7.**
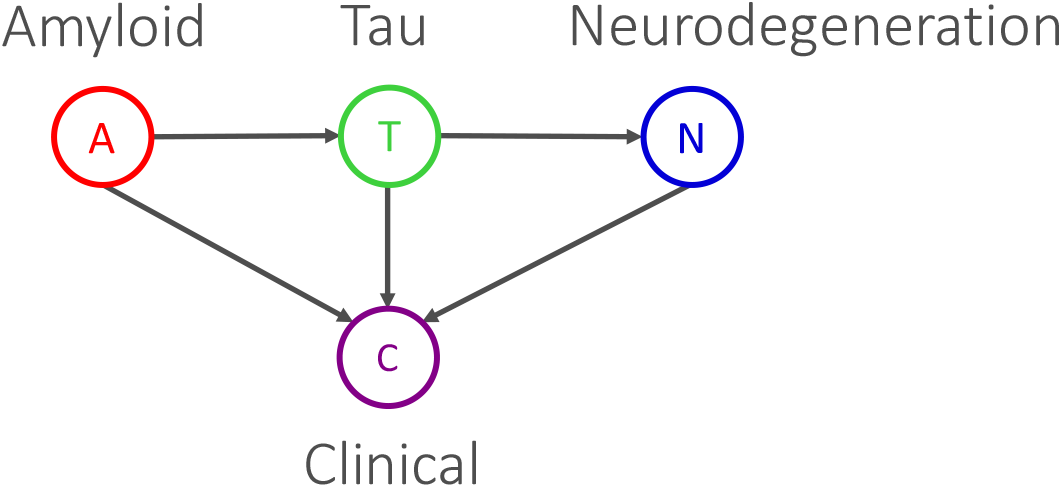
The graph represents the relationships encoded by SimulAD to predict longitudinal multimodal trajectories in the training cohort.

#### 4.2.4 Intervention Model

Drug intervention is modelled by augmenting Eq. 1 with an additive clearance term:

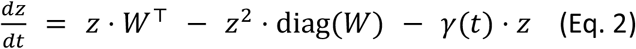

where *γ* is a piecewise-constant clearance rate applied to the AV45 latent dimension. The piecewise structure mirrors published dosing schedules: an initial active phase (months 0–6 except 0–12 for gantenerumab), a maintenance phase (months 6–18 for all trials except 12–27 for gantenerumab), and cessation (*γ* = 0) at or after the primary endpoint. The *γ* values were calibrated to reproduce published centiloid reductions (Section 2.2, Table 1). Owing to the cascade structure of SimulAD (Section 4.2.3), reducing AV45 propagates downstream effects on tau, MRI atrophy, and CDR-SB according to the learned interaction coefficients. The piecewise *γ* values were calibrated iteratively by scaling *γ* by the ratio k = %CL_real / %CL_sim at each iteration, where %CL_real is the published fractional centiloid reduction at the primary endpoint and %CL_sim is the corresponding simulated reduction. Two scaling iterations were performed; final values are reported in Table 1.

### 4.3 Simulations

Population simulation proceeds in three stages: (i) trial-specific baseline calibration via latent distribution centre optimisation, (ii) ODE integration to generate full trajectories, and (iii) biomarker decoding (pseudo-code provided in Supplementary Section S3). The code, model parameters and simulations will be made available upon acceptance of this work.

#### 4.3.1 Baseline calibration

A Gaussian latent distribution is first fitted to the encoded baseline latent states of the training cohort. A trial-specific latent centre and covariance scale are then optimised by minimising:

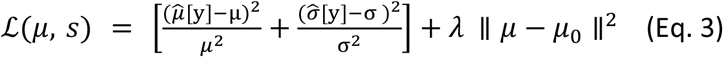

where (μ, σ) are the target CDR-SB mean and SD from the published trial placebo baseline (3; 4; 5), and 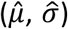 are Monte Carlo estimates of mean and SD of predicted CDR-SB computed from N = 500 samples drawn from the latent distribution ℱ(*μ*_*z*_, Σ_*z*_), with *μ*_*z*_ and Σ_*z*_ respectively the latent mean and covariance matrix of the training cohort. To reflect eligibility criteria, the training cohorts were initialised from ADNI clinical subgroups: MCI converters to AD (MCI→AD) for TRAILBLAZER, CLARITY, EMERGE, and ENGAGE; MCI converters and mild AD for GRADUATE I and GRADUATE II (Bateman et al., 2023).

The parameter *λ* = 0.1 is an L2 regularisation weight. Optimisation used Adam (lr = 5 × 10^−3^, 1000 iterations) (22).

#### 4.3.2 ODE Integration

Starting from the optimised GMM, N ∈ {50, 100, 200, 300, 400, 500, 800} subjects per arm are independently sampled for each simulation repetition. Each trajectory *z*(t) is integrated forward in time from trial-specific latent centres (Section 4.3.1), with t = 0 representing simulated enrolment and with target trial end point month (27 months for GRADUATE I and II or 18 months for the other trials). The placebo arm follows Eq. 1; the treated arm follows Eq. 2 with the trial-specific piecewise *γ*(t) schedule.

#### 4.3.3 Biomarker decoding

CDR-SB is obtained from the CLINIC decoder output while centiloid from the AV45 decoder output, both denormalised using per-modality mean and SD from the training cohort. Simulated visits are aligned with published trial schedules (every 3 months to the primary endpoint: 18 months for TRAILBLAZER-ALZ2, CLARITY AD, EMERGE, and ENGAGE; 27 months for GRADUATE I and GRADUATE II).

### 4.4. Model training

SimulAD was trained by maximising the evidence lower bound (ELBO):

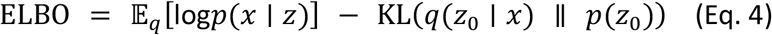

where p(*z*_0_) = N(0, I) is the standard Gaussian prior on the initial latent state and p(x | z) is the Gaussian reconstruction likelihood with per-biomarker learnable variance. The expectation is estimated via the reparameterisation trick; missing biomarker values are excluded from the likelihood. Optimisation used Adam (22) with lr = 1 × 10^−4^, run for 50 000 epochs with gradient clipping at max norm 1.0. Training data were batched into nine time-windows (e.g., [0, 3], [0, 6], [−3, 0], [−6, 0] years) to balance subjects across follow-up lengths. The model was implemented in PyTorch; ODE integration used torchdiffeq. Training code is available at https://gitlab.inria.fr/epione_ML/simulad_v2.

### 4.5 Statistical Analysis

#### 4.5.1 Linear Mixed Effect Models for Repeated Measures (LMM)

Treatment effects on CDR-SB were estimated using a linear mixed model for repeated measures (LMM), consistent with the primary analysis plans of the original trials. For each simulated cohort the following linear mixed model was fitted (statsmodels, REML, L-BFGS with Powell fallback) (23):

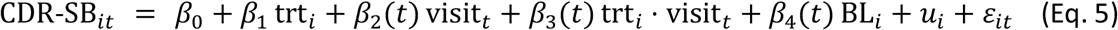

where trt_*i*_ ∈ {0, 1} is the treatment indicator, visit_*t*_ is a categorical time variable, BL_*i*_ is the subject’s CDR-SB at baseline, *u*_*i*_ ∼ η(0, *τ*^2^) is a subject-level random intercept, and *ε*_it_ is the residual error. The treatment effect at the primary endpoint T was estimated as the Wald contrast *β*_1_ + *β*_3_(T), with SE derived from the model covariance matrix. Statistical significance was declared at *α* = 0.05 (two-tailed).

#### 4.5.2 Empirical Power Analysis

Statistical power was estimated empirically by running 500 independent simulation repetitions at each of seven sample sizes per arm: N ∈ {50, 100, 200, 300, 400, 500, 800}. For each repetition, two independent synthetic cohorts of N subjects per arm were generated, trajectories were simulated, and the LMM was fitted to CDR-SB at all scheduled visits. Power at each N was estimated as the proportion of repetitions yielding p < 0.05. The minimum sample size for 80% power (N_80_) was estimated by linear interpolation of the empirical power curve.

#### 4.5.3 Effect size

The simulated treatment effect is reported as the LMM estimate 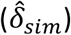 averaged across 500 repetitions ± 1 SD, in CDR-SB points. Real-world treatment effects were taken from the published primary analyses: −0.70 (TRAILBLAZER-ALZ2), −0.45 (CLARITY AD), −0.39 (EMERGE), +0.03 (ENGAGE), −0.31 (GRADUATE I), and −0.19 (GRADUATE II). Trials were classified as significant if p < 0.05 in their published primary analysis.

#### 4.5.4 Follow-up Calibration: Effect-size, Variance, Power and Simulations

SimulAD’s raw LMM estimate 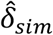 and simulated pooled standard deviation 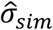 are systematically lower than published trial values. Underestimation is likely due to a number of concomitant causes, including SimulAD statistical compression and the ADNI cohort homogeneity causing out-of-distribution ODE dynamics and placebo cohort underestimation (driving 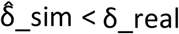), and lower CDR-SB inter-subject variability of ADNI than the broader, more heterogeneous populations enrolled in phase III trials (driving 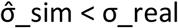).

To account for this issue, the following independent leave-one-out (LOO) correction multipliers are estimated for each target trial t from the remaining calibration trials C = {TRAILBLAZER, CLARITY, EMERGE, GRADUATE I, GRADUATE II}. ENGAGE was excluded from the set of trials C because its real effect is near zero.

##### Effect multiplier

For each trial t ∈ C, the per-trial ratio 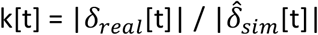 quantifies how much the SimulAD simulated LMM effect 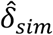 underestimates the published real LMM treatment effect *δ*_*real*_ at the primary endpoint. Because of the linear relationship observed between k and absolute amyloid reduction rate ΔCL/trial duration (k = 0.5 + 0.48 × ΔCL/t, R^2^ = 0.85, Supplementary Section S6) we derived an automated calibration procedure to estimate the trial-specific ratio 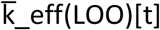 from the other trials through a linear model prediction via leave-one-out (LOO). For ENGAGE, k is predicted from the full five-trial fit (out-of-sample, since ENGAGE ∉ C).

##### Variance multiplier

For each trial t (all six, including ENGAGE), we computed the per-trial SD ratio 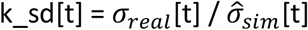, where *σ*_*real*_[t] is the pooled standard deviation of CDR-SB change from baseline at the primary endpoint reported in the published trial (derived from the trial’s own LMM analysis and pooled across treated and placebo arms), and 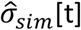 is the corresponding SimulAD simulated pooled SD. ENGAGE is included in the SD calibration set because the SD ratio is independent of treatment direction. The LOO SD multiplier for target trial t is 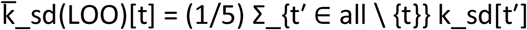, averaging over the five remaining trials (cross-trial mean ≈ 1.81, range 1.65–2.11).

##### Calibrated N_80_

Three calibration strategies are used, differing in what target-trial data are used as input and, consequently, what assumptions are required.

i. **Fully prospective (primary strategy)**. With this strategy no real target endpoint data from target trial t are required. The calibrated treatment effect 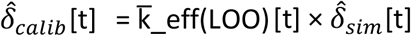, and calibrated pooled SD 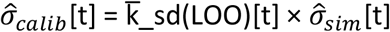 are both estimated using only LOO multipliers. Sample size analysis is performed by computing 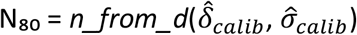, where *n_from_d*(-) is the standard two-sample t-test power formula at α = 0.05 (two-sided), power = 0.80, equal allocation.
ii. **Semi-prospective (SD benchmark)**. The calibrated effect 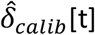 is retained, but 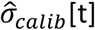 is replaced by the published real SD *σ*_*real*_[t] from the target trial. Sample size analysis is performed by computing 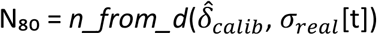. This strategy is not prospective with respect to the SD (it requires knowing *σ*_*real*_[t] in advance), but it isolates the contribution of effect-size miscalibration from SD miscalibration, providing an upper bound on what would be achievable with a perfect variance model.
iii. **Oracle (retrospective reference)**. Sample size is computed with N_80_ = *n_from_d*(*δ*_*real*_, *σ*_*real*_), using the published real effect and real SD. This is the minimum N that would achieve 80% power with perfect knowledge of the real trial outcomes.

##### Simulations

To apply the calibration to the raw SimulAD predictions, for each trial t we construct three calibrated predictions of the mean treated arm follow-up mean at time T, μ_treated_(T)[t], all using the same calibrated treatment effect 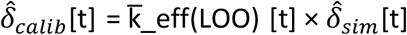, but differing in the placebo follow-up anchor. Similarly, in all simulations the treated follow-up error bar uses 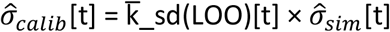, the same prospective calibrated pooled SD as used in the N_80_ calculation (Section 4.5.4); the placebo follow-up SD is instead trial-specific and detailed below:

i. **LOO-calibrated placebo (primary prediction)**. This prediction represents the prospective setting in which prior knowledge provides a prior estimate of placebo CDR-SB progression, for example calibrated from a Phase 2 study or from other trials (as in the LOO setting detailed above). In this setting, 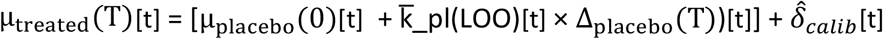, where Δ_placebo_(T)[t] = (μ_placebo_(T) − μ_placebo_(0))[t] is the simulated placebo CDR-SB change from baseline to the primary endpoint for trial t, and 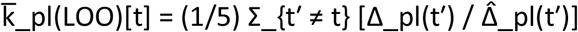 is the LOO mean of the per-trial placebo-progression ratio for trial t (published CDR-SB change divided by simulated CDR-SB change, analogous to 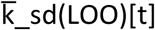 for the variance multiplier) (Supplementary Figure S4). The placebo follow-up SD is 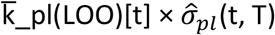, where 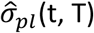 is the simulated placebo follow-up standard deviation.
ii. **Pure SimulAD**. 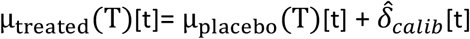, where μ_placebo_(T)[t] is the SimulAD-simulated placebo follow-up CDR-SB mean (raw simulation output, no drug effect applied) (Figure 4 and Supplementary Figure S4). The placebo follow-up SD is 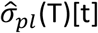, the raw simulated placebo follow-up standard deviation.
iii. **Real-anchored**. Here we compute 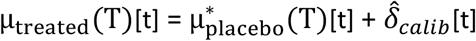, where 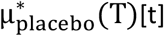 is the published placebo follow-up CDR-SB mean. By anchoring to the real placebo trajectory this series isolates the accuracy of treatment-effect calibration from any errors in the simulated placebo progression (Supplementary Figure S4). The placebo follow-up SD is *σ*_*pl*_(T)[t], the published placebo follow-up standard deviation.

A detailed description of estimated calibration coefficients and related simulation results across trials id provided in Supplementary Section S4.

### 4.6 Comparison Baselines

Three reference approaches were compared against SimulAD to assess the added value of mechanistic kinetic simulation over standard statistical methods trained on observational data.

#### Baseline 1 (Parametric power formula)

*N*_80_ was computed analytically from SimulAD’s LMM treatment effect estimate 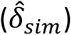 and pooled standard deviation 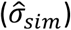 using the standard two-sample t-test formula 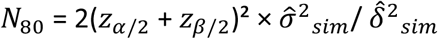, with *α* = 0.05 (two-sided) and 1 − *β* = 0.80.

#### Baseline 2 (Ridge regression)

A Ridge regression pipeline (scikit-learn StandardScaler + Ridge Regression, default regularisation (24)) was trained on ADNI visit pairs constructed from consecutive post-baseline visits 6-30 months apart for which both centiloid (AV45) and CDR-SB were non-missing (n = 126 pairs, 187 subjects). Features were centiloid and CDR-SB at the earlier visit, centiloid at the later visit, and elapsed time (months); the target was CDR-SB at the later visit. Predictive accuracy was evaluated by 5-fold cross-validation. Trial CDR-SB treatment effects were estimated by predicting CDR-SB at the primary endpoint for the treated arm, where centiloid at the later visit was computed as the difference between baseline and drug-specific reduction using published baseline CDR-SB and centiloid means. For the placebo arm, centiloid at later visit was estimated as baseline + 0.21 × endpoint months, based on the median ADNI natural accumulation rate.

#### Baseline 3 (Gradient boosting)

A GradientBoostingRegressor (n_estimators = 200, max_depth = 3, learning_rate = 0.05, scikit-learn (24)) was trained and applied identically to Baseline 2.

### 4.7 Multimodal cascade analysis

Statistical association between amyloid reduction and downstream dynamics of biomarker changes was assessed by correlation analysis, and by inspecting the coupling ODE weights learned by SimulAD. To quantify the predicted downstream effects of amyloid clearance on tau pathology and structural MRI at the individual level, we extracted biomarker trajectories from the same SimulAD simulations used for CDR-SB analysis (Section 4.3). For each subject i and each arm (treated, placebo), we computed individual change scores at the primary endpoint: ΔAV45_i_ = AV45(BL) − AV45(endpoint), ΔTAU_i_ = TAU(BL) − TAU(endpoint), and ΔVol_i_ = MRI(BL) − MRI(endpoint), where positive values denote a reduction. Statistical association between amyloid change and each downstream biomarker was assessed by Pearson r and Spearman ρ of ΔAV45 vs ΔTAU and ΔAV45 vs ΔVol, computed separately within the treated and placebo arms. To remove between-trial mean-level differences before pooling, change scores were z-scored within each trial prior to aggregation across the six trials. Analyses were conducted on N = 3,200 subjects per arm (8 simulation repetitions × 400 subjects).

## Data Availability

All data produced in the present study are available upon reasonable request to the authors

## 5. Acknowledgements

Data collection and sharing for the Alzheimer’s Disease Neuroimaging Initiative (ADNI) is funded by the National Institute on Aging (National Institutes of Health Grant U19AG024904). The grantee organization is the Northern California Institute for Research and Education. In the past, ADNI has also received funding from the National Institute of Biomedical Imaging and Bioengineering, the Canadian Institutes of Health Research, and private sector contributions through the Foundation for the National Institutes of Health

(FNIH) including generous contributions from the following: AbbVie, Alzheimer’s Association; Alzheimer’s Drug Discovery Foundation; Araclon Biotech; BioClinica, Inc.; Biogen; Bristol-Myers Squibb Company; CereSpir, Inc.; Cogstate; Eisai Inc.; Elan Pharmaceuticals, Inc.; Eli Lilly and Company; EuroImmun; F. Hoffmann-La Roche Ltd and its affiliated company Genentech, Inc.; Fujirebio; GE Healthcare; IXICO Ltd.; Janssen Alzheimer Immunotherapy Research & Development, LLC.; Johnson & Johnson Pharmaceutical Research & Development LLC.; Lumosity; Lundbeck; Merck & Co., Inc.; Meso Scale Diagnostics, LLC.; NeuroRx Research; Neurotrack Technologies; Novartis Pharmaceuticals Corporation; Pfizer Inc.; Piramal Imaging; Servier; Takeda Pharmaceutical Company; and Transition Therapeutics.

## 6. Supplementary Material

### S1. ADNI Training Cohort: Baseline Summary Statistics

SimulAD was trained on 351 amyloid-positive participants from the Alzheimer’s Disease Neuroimaging Initiative (ADNI) with complete multi-modal imaging and cognitive data at baseline (see Methods). Participants were stratified into five longitudinal trajectory classes: clinically normal (NL, n = 174), NL converters to MCI (NL → MCI, n = 19), MCI (n = 81), MCI converters to AD (MCI → AD, n = 38), and clinically diagnosed AD (n = 39). Longitudinal follow-up yielded a total of 1,256 visits across 351 subjects (mean 3.6 visits/subject; range 3.3–5.6 across stages), providing the repeated-measures trajectories used to train the latent ODE dynamics.

Table S1 reports mean (SD) values for all biomarkers and clinical scores at baseline, stratified by stage. The cohort exhibits the expected gradient of disease severity across stages: amyloid burden (measured by centiloid, CL) increases from 38.5 (32.5) CL in NL to 100.0 (37.3) CL in AD; intra-cranial volume (ICV) normalized hippocampal volume decreases from 5.19 (0.60) × 10^3^ to 3.87 (0.83) × 10^3^; entorhinal tau SUVR increases from 1.19 (0.15) to 1.61 (0.24); and CDR-SB increases from 0.06 (0.21) in NL to 4.17 (1.71) in AD. The distribution of biomarker values across stages, illustrated in Figure S1, confirms that the training cohort spans the full spectrum of amyloid-positive disease, from preclinical to dementia, as required for unbiased modelling of disease progression.

**Table S1.** Baseline characteristics of the ADNI training cohort (N = 351 amyloid-positive participants), stratified by longitudinal trajectory class. Values are mean (SD). NL = Clinically Normal; MCI = Mild Cognitive Impairment; AD = Alzheimer’s Disease. SUVR = Standardised Uptake Value Ratio (flortaucipir tau PET or AV45 amyloid PET). MRI volumes are ICV-normalised.

| Biomarker | NL | NL to MCI | MCI | MCI to AD | AD | Total |
| --- | --- | --- | --- | --- | --- | --- |
| **N (subjects)** | **174** | **19** | **81** | **38** | **39** | **351** |
| **Amyloid PET (AV45)** |  |  |  |  |  |  |
| Centiloid | 38.5 (32.5) | 64.0 (43.5) | 65.2 (46.6) | 86.5 (34.8) | 100.0 (37.3) | 58.1 (43.4) |
| Inf. parietal (SUVR) | 1.24 (0.20) | 1.44 (0.27) | 1.38 (0.27) | 1.51 (0.21) | 1.61 (0.25) | 1.36 (0.26) |
| Inf. temporal (SUVR) | 1.19 (0.18) | 1.36 (0.27) | 1.32 (0.27) | 1.47 (0.21) | 1.59 (0.24) | 1.30 (0.26) |
| Mid. temporal (SUVR) | 1.15 (0.19) | 1.34 (0.26) | 1.29 (0.26) | 1.43 (0.21) | 1.54 (0.24) | 1.27 (0.26) |
| Post. cingulate (SUVR) | 1.30 (0.22) | 1.45 (0.27) | 1.47 (0.28) | 1.58 (0.20) | 1.67 (0.26) | 1.42 (0.27) |
| Precuneus (SUVR) | 1.30 (0.23) | 1.49 (0.29) | 1.46 (0.29) | 1.63 (0.21) | 1.72 (0.27) | 1.43 (0.29) |
| Sup. temporal (SUVR) | 1.12 (0.17) | 1.25 (0.23) | 1.24 (0.24) | 1.35 (0.18) | 1.45 (0.21) | 1.22 (0.23) |
| Supramarginal (SUVR) | 1.21 (0.18) | 1.35 (0.24) | 1.34 (0.27) | 1.46 (0.18) | 1.55 (0.23) | 1.32 (0.24) |
| **MRI volumes (ICV-normalised)** |  |  |  |  |  |  |
| Ventricles ($\times 10^3$) | 22.00 (10.70) | 33.83 (19.96) | 27.08 (11.81) | 32.45 (11.59) | 34.69 (14.02) | 26.36 (12.99) |
| Hippocampus ($\times 10^3$) | 5.19 (0.60) | 4.94 (0.71) | 4.70 (0.62) | 4.17 (0.75) | 3.87 (0.83) | 4.80 (0.80) |
| Whole brain | 0.721 (0.041) | 0.716 (0.042) | 0.704 (0.040) | 0.681 (0.062) | 0.661 (0.097) | 0.706 (0.056) |
| Fusiform ($\times 10^3$) | 12.95 (1.29) | 12.43 (1.32) | 12.54 (1.59) | 11.34 (2.14) | 11.06 (1.85) | 12.44 (1.68) |
| Mid. temporal ($\times 10^3$) | 14.47 (1.59) | 13.95 (1.26) | 13.65 (1.76) | 12.56 (1.87) | 12.09 (2.03) | 13.78 (1.89) |
| **Tau PET (flortaucipir)** |  |  |  |  |  |  |
| Entorhinal (SUVR) | 1.19 (0.15) | 1.30 (0.20) | 1.40 (0.32) | 1.60 (0.31) | 1.61 (0.24) | 1.33 (0.28) |
| Banks STS (SUVR) | 1.22 (0.11) | 1.32 (0.28) | 1.35 (0.31) | 1.65 (0.59) | 1.80 (0.68) | 1.37 (0.40) |
| Fusiform (SUVR) | 1.21 (0.10) | 1.30 (0.19) | 1.36 (0.28) | 1.65 (0.45) | 1.70 (0.41) | 1.35 (0.31) |
| Parahippocampal (SUVR) | 1.13 (0.11) | 1.23 (0.18) | 1.27 (0.24) | 1.44 (0.26) | 1.47 (0.24) | 1.24 (0.22) |
| Amygdala (SUVR) | 1.25 (0.15) | 1.34 (0.23) | 1.46 (0.33) | 1.67 (0.32) | 1.66 (0.28) | 1.39 (0.29) |
| Hippocampus (SUVR) | 1.27 (0.14) | 1.27 (0.20) | 1.36 (0.24) | 1.45 (0.20) | 1.44 (0.22) | 1.33 (0.20) |
| **Clinical assessments** |  |  |  |  |  |  |
| CDR-SB | 0.06 (0.21) | 0.42 (0.63) | 1.46 (1.25) | 4.12 (3.27) | 4.17 (1.71) | 1.30 (2.11) |
| MMSE | 29.2 (1.0) | 28.5 (1.9) | 27.6 (2.1) | 24.6 (4.3) | 22.8 (2.8) | 27.6 (3.1) |
| ADAS-11 | 5.1 (2.8) | 6.8 (3.9) | 9.2 (3.7) | 16.4 (8.5) | 20.2 (5.8) | 9.1 (6.8) |
| ADAS-13 | 8.0 (4.1) | 11.3 (6.5) | 15.2 (5.9) | 25.7 (10.8) | 31.1 (7.3) | 14.3 (10.1) |
| ADAS-Q4 | 2.4 (1.7) | 3.7 (2.6) | 5.2 (2.4) | 7.7 (2.4) | 9.1 (1.4) | 4.5 (3.1) |
| FAQ | 0.2 (0.6) | 0.7 (1.4) | 3.1 (4.2) | 11.0 (8.0) | 12.9 (6.6) | 3.5 (6.1) |
| MoCA | 26.6 (2.6) | 23.5 (3.3) | 23.0 (3.5) | 19.2 (5.0) | 16.5 (4.5) | 23.7 (4.9) |
| RAVLT Immediate | 47.6 (10.3) | 38.5 (8.1) | 36.1 (10.4) | 27.1 (10.0) | 22.4 (7.1) | 39.5 (13.5) |
| RAVLT Learning | 6.4 (2.2) | 4.8 (2.5) | 4.0 (2.6) | 2.4 (2.0) | 2.2 (1.7) | 4.9 (2.8) |
| RAVLT Forgetting | 3.8 (2.9) | 4.1 (3.6) | 4.9 (2.4) | 4.0 (6.7) | 4.9 (2.3) | 4.2 (3.4) |
| Logical Memory (LDEL) | 13.8 (3.5) | 11.0 (3.7) | 7.8 (4.4) | 4.0 (4.4) | 1.5 (2.1) | 9.8 (5.8) |

**Figure S1.**
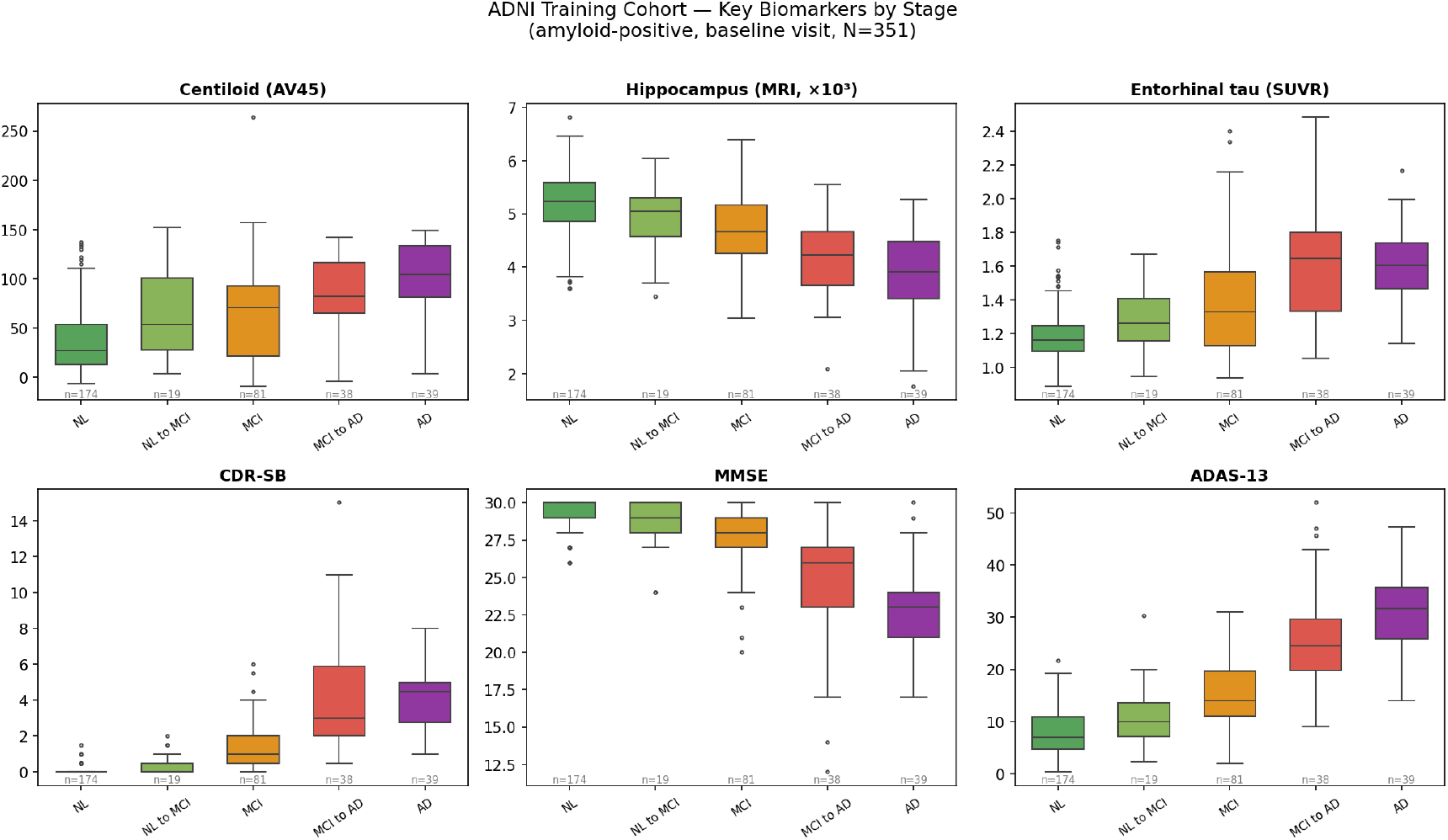
Baseline biomarker distributions by longitudinal trajectory class (N = 351 amyloid-positive ADNI participants). Box plots show median (IQR) with individual data points overlaid. From left to right within each panel: NL (clinically normal), NL → MCI (converters), MCI, MCI → AD (converters), AD. Panels show, clockwise from top left: amyloid burden (centiloid, AV45 PET), hippocampal volume (MRI, ICV-normalised), entorhinal cortex tau (flortaucipir SUVR), CDR-SB, and MMSE.

### S2. SimulAD Disease Dynamics: Biomarker Velocity Fields

The latent ODE at the core of SimulAD defines a continuous-time vector field over the joint biomarker space. To characterise the model’s learned disease dynamics, we computed the instantaneous rate of change of each biomarker as a function of the current disease state, evaluated at the baseline observations of all 351 training subjects. The rate of change of each biomarker b at state z is given by the decoder-projected latent velocity: 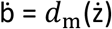, where ż = f(z; W) is the latent ODE right-hand side (Eq. 1 in the main text) and *d*_m_ is the corresponding decoder. Figure S2 shows the predicted velocity fields for amyloid burden (centiloid, d(CL)/dt) and cognitive decline (CDR-SB, d(CDR-SB)/dt) as a function of the joint (centiloid, CDR-SB) state. Amyloid accumulation rates ranged from 1.88 to 4.77 CL/year across the observed subject distribution, with the highest rates concentrated in subjects with elevated amyloid but preserved cognition, consistent with the preclinical accumulation phase. CDR-SB progression rates ranged from −0.03 to 1.21 points/year; the negative-to-zero values reflect subjects with apparent cognitive fluctuation near the floor of the scale. The velocity field structure confirms that SimulAD captures the amyloid-driven acceleration of cognitive decline: CDR-SB progression rates increase substantially once amyloid burden exceeds approximately 75 CL, consistent with the amyloid threshold hypothesis.

**Figure S2.**
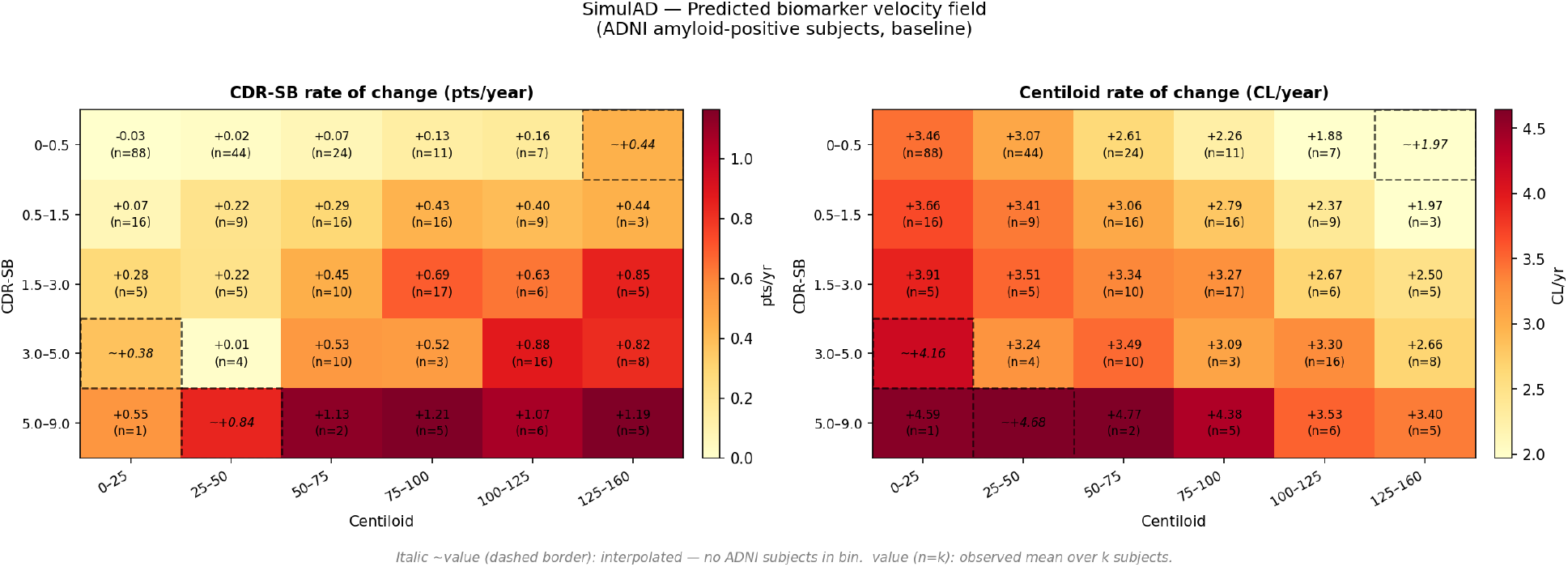
SimulAD velocity field for amyloid burden (d(CL)/dt, left) and cognitive decline (d(CDR-SB)/dt, right) as a function of the joint (centiloid, CDR-SB) state. Each cell shows the mean predicted instantaneous rate of change for subjects falling within the corresponding (centiloid, CDR-SB) bin. Colour scale is in CL/year (left) and points/year (right). Cells with dashed contours indicate extrapolated values with no representative individuals in the training cohort.

Figure S3 extends the velocity field analysis to tau PET and structural MRI. Tau accumulation rates (entorhinal SUVR/year) increase monotonically with both amyloid burden and cognitive stage, consistent with the downstream position of tau pathology in the proposed SimulAD causal pathway. Hippocampal atrophy rates (× 10^3^/year) likewise scale with disease severity, with the highest atrophy rates predicted in subjects with advanced cognitive impairment, mirroring the tau-mediated neurodegeneration cascade.

**Figure S3.**
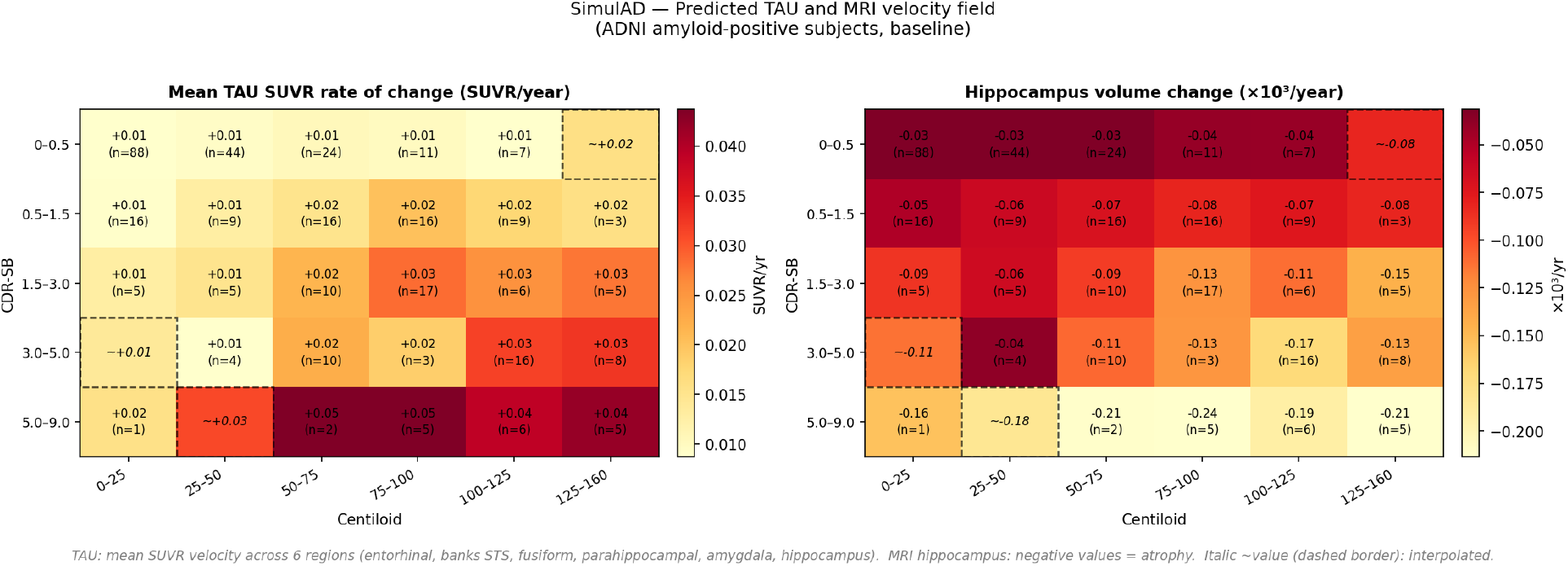
SimulAD velocity field for tau burden (d(entorhinal SUVR)/dt, left) and hippocampal atrophy (d(hippocampus)/dt, right) as a function of the joint (centiloid, CDR-SB) state. Layout as in Figure S2. Cells with dashed contours indicate extrapolated values with no representative individuals in the training cohort.

### S3. SimulAD Trial simulation pseudo-code

#### Algorithm 1.

SimulAD Trial Simulation

**Algorithm 1**. SimulAD trial simulation procedure. Given a target population specification (baseline centiloid and CDR-SB) and drug dynamics *γ*(t), the procedure produces simulated CDR-SB trajectories for placebo and treated arms over R repetitions, and returns the mean LMM treatment effect, empirical power at arm size N, and the sample size required for 80% power (N_80_). Equation references correspond to the main text; Figure S6 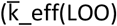 vs amyloid clearance regression) is referenced in the supplementary.

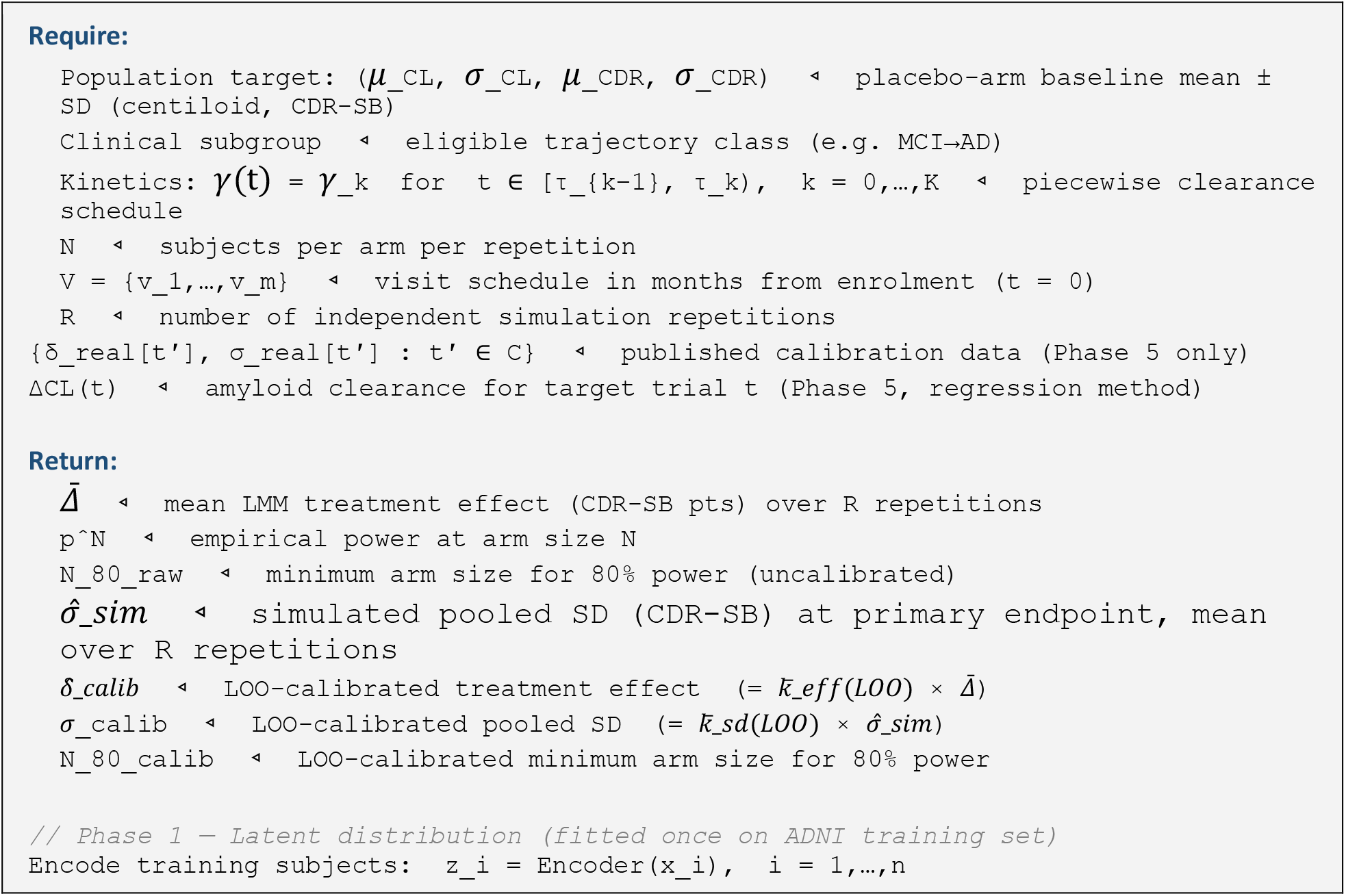
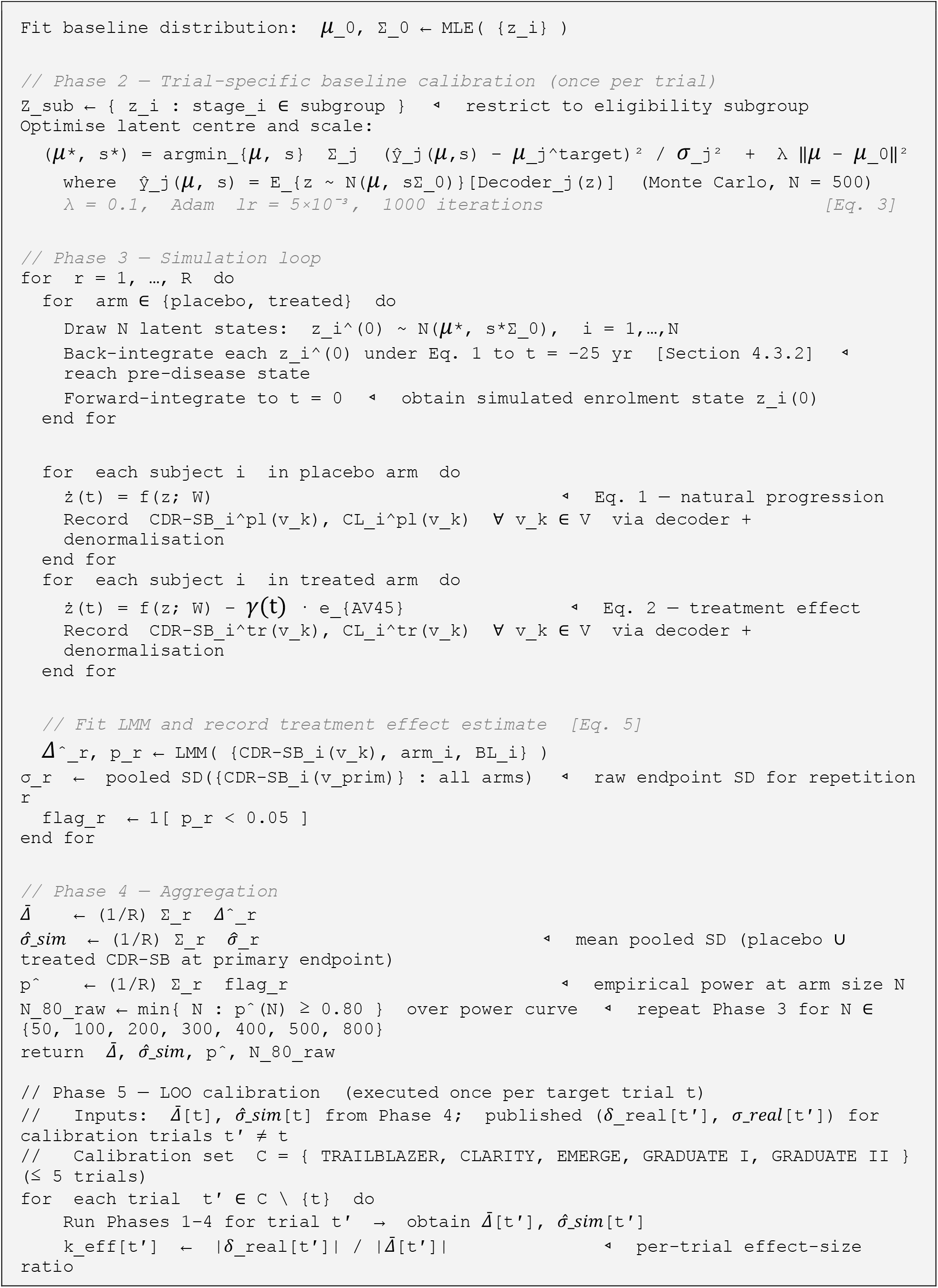
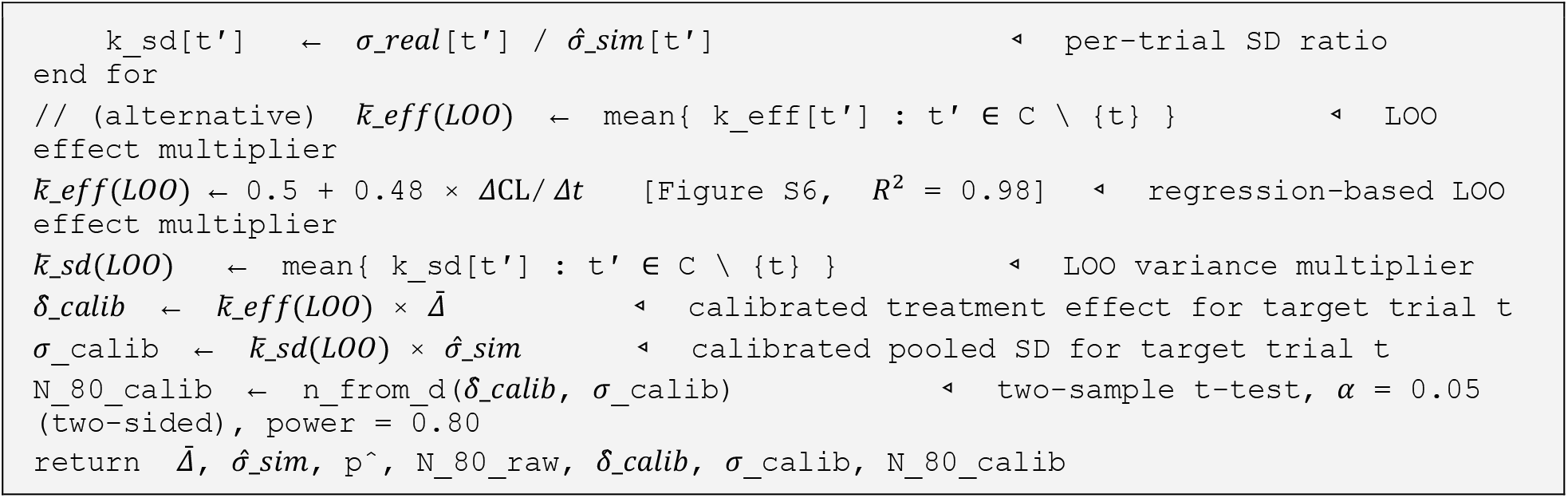

### S4. Calibrations comparisons and simulation results

Raw SimulAD simulations reproduce the qualitative direction of treatment effects across all six trials but systematically underestimate both the LMM treatment effect and the pooled CDR-SB standard deviation. Table S4a summarises the raw simulation outputs alongside published values.

Table S4b details the full set of calibration coefficients for all six trials. The mean ratio of real-to-simulated effect size 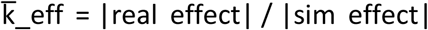 across the five trials with a positive treatment effect (all except ENGAGE, whose published effect is near zero) ranges from 0.97 (GRADUATE II) to 2.71 (TRAILBLAZER). The SD ratio (k_sd = *σ*_*real* / *σ*_*sim*) is more stable: range 1.64–2.10. Both underestimations reflect the gap between ADNI’s observational cohort and phase III trial populations.

The per-trial effect multiplier 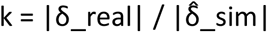 varies substantially across trials (range 0.97– 2.71), with the largest value for TRAILBLAZER. The LOO estimate 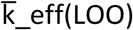 yields values in the range 1.35–3.37. ENGAGE is excluded from the calibration set C (its real effect is near zero and of uncertain sign) and its 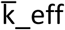 is predicted out-of-sample from the full five-trial fit. The SD multiplier 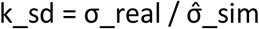 is more stable across trials (range 1.64–2.10), reflecting a systematic and near-constant underestimation of CDR-SB inter-subject variability by the ADNI-trained model. The LOO SD multiplier 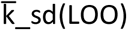 is particularly stable (range 1.74–1.83), confirming that the five-trial leave-one-out mean is a robust prospective estimate. The resulting calibrated effects 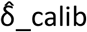 range from −0.25 (GRADUATE I) to −0.86 (TRAILBLAZER), closely bracketing the published real effects (range −0.19 to −0.70), with a mean absolute error of 0.09 CDR-SB points across the five calibration trials. The calibrated SDs 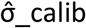 (range 2.46–2.97) are similarly close to published σ_real (range 2.20–3.22), confirming that the LOO multipliers recover realistic variance estimates without requiring any target-trial data. Figure S4 shows the resulting CDR-SB predictions for all the calibration methods. Predictions results are coherent across approaches for both placebo and treated arms.

**Figure S4a.**
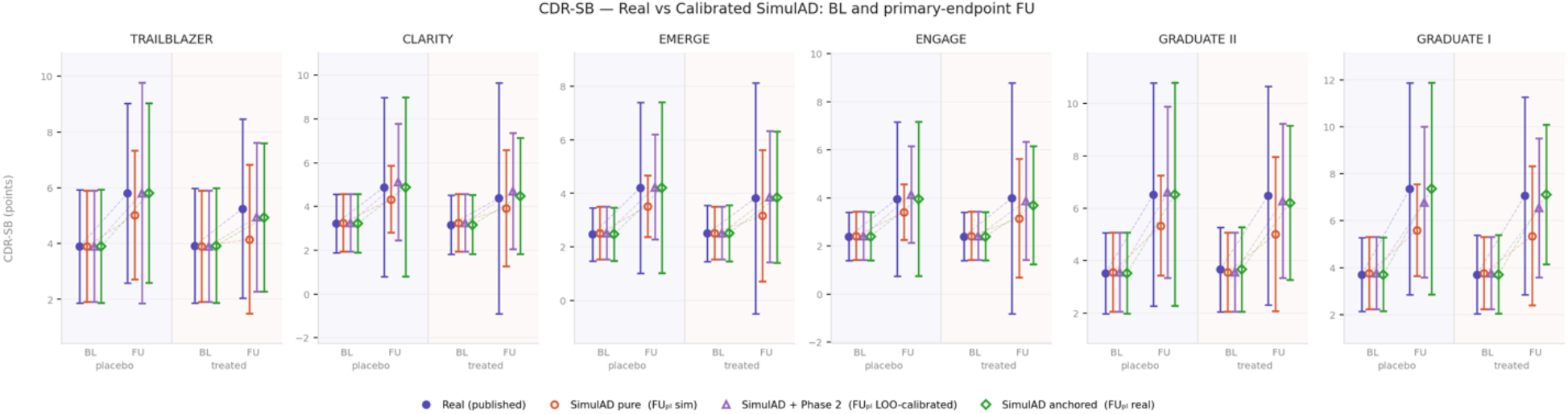
Head-to-head comparison of CDR-SB predictions across three SimulAD reconstruction scenarios versus published trial values. For each trial, mean CDR-SB ± 1 SD is shown for placebo and treated arms at baseline and primary-endpoint follow-up under: raw simulation (red circles), LOO prospective calibrated (purple triangles), and real-anchored (green diamonds) (Methods Section 4.5.4). Published values are shown as blue filled circles.

**Table S4a.**
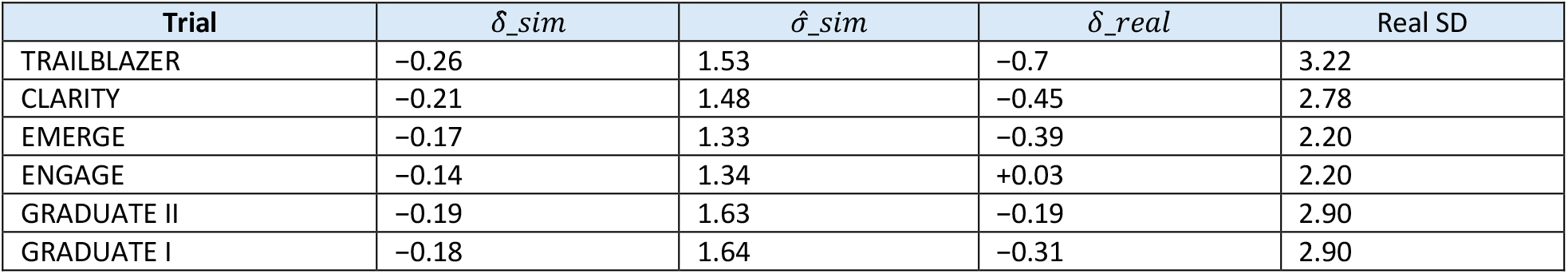
Uncalibrated CDR-SB simulation outputs vs published trial values. 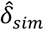: raw LMM effect (mean ± SD, 500 repetitions). *σ*_*sim*_: simulated pooled SD.

**Table S4b.** Calibration coefficients for all six trials. δ_real: published LMM treatment effect (CDR-SB points).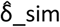: SimulAD simulated LMM effect (mean over 500 repetitions at N = 800).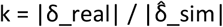: per-trial effect ratio (n/a for ENGAGE, excluded from calibration set C).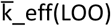: LOO-predicted effect multiplier from k∼ΔCL linear regression (LOO for calibration trials; out-of-sample for ENGAGE).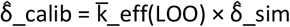. σ_real: published pooled CDR-SB SD at primary endpoint.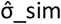: SimulAD simulated pooled SD.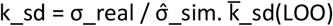: LOO mean SD multiplier (mean of k_sd over the five other trials).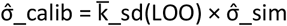.

| Trial | Effect-size coefficients |  |  |  |  | Variance coefficients |  |  |  |  |
| --- | --- | --- | --- | --- | --- | --- | --- | --- | --- | --- |
| | $\delta_{real}$ | $\hat{\delta}_{sim}$ | k | $\bar{k}_{eff}(LOO)$ | $\hat{\delta}_{calib}$ | $\sigma_{real}$ | $\hat{\sigma}_{sim}$ | $k_{sd}$ | $\bar{k}_{sd}(LOO)$ | $\hat{\sigma}_{calib}$ |
| TRAILBLAZER | -0.70 | -0.26 | 2.71 | 3.37 (LOO) | -0.86 | 3.22 | 1.53 | 2.10 | 1.74 | 2.66 |
| CLARITY | -0.45 | -0.21 | 2.17 | 1.96 (LOO) | -0.40 | 2.78 | 1.48 | 1.87 | 1.79 | 2.65 |
| EMERGE | -0.39 | -0.17 | 2.26 | 2.09 (LOO) | -0.36 | 2.20 | 1.35 | 1.65 | 1.83 | 2.45 |
| ENGAGE | +0.03 | -0.14 | - | 1.95 (OOS) | -0.27 | 2.20 | 1.34 | 1.64 | 1.83 | 2.46 |
| GRADUATE II | -0.19 | -0.19 | 0.97 | 1.64 (LOO) | -0.32 | 2.90 | 1.63 | 1.78 | 1.81 | 2.94 |
| GRADUATE I | -0.31 | -0.18 | 1.69 | 1.35 (LOO) | -0.25 | 2.90 | 1.64 | 1.77 | 1.81 | 2.97 |

### S5. Statistical power analysis across calibrated and uncalibrated simulations

Table S5 summarises N_80_ and power at the actually enrolled N for each trial under four strategies: Oracle (published real effect and SD), fully prospective 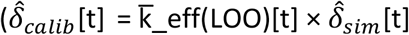 and 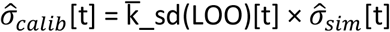, no target endpoint data from the trial required), semi-prospective (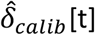 with published real SD *σ*_*real*_), and raw uncalibrated SimulAD 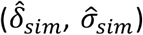. Figure S5 shows empirical power curves for the raw and fully prospective strategies. At the actually enrolled N, the fully prospective strategy correctly classifies TRAILBLAZER as well-powered (100%) and both GRADUATE trials as under-powered (≤50%), consistent with published outcomes. EMERGE is borderline (65%), in line with its modest significance (p = 0.01). ENGAGE is correctly identified as under-powered under all strategies. Raw simulations overestimate power for GRADUATE II (53% vs oracle 18%) and underestimate for EMERGE (55% vs oracle 84%).

**Figure S5.**
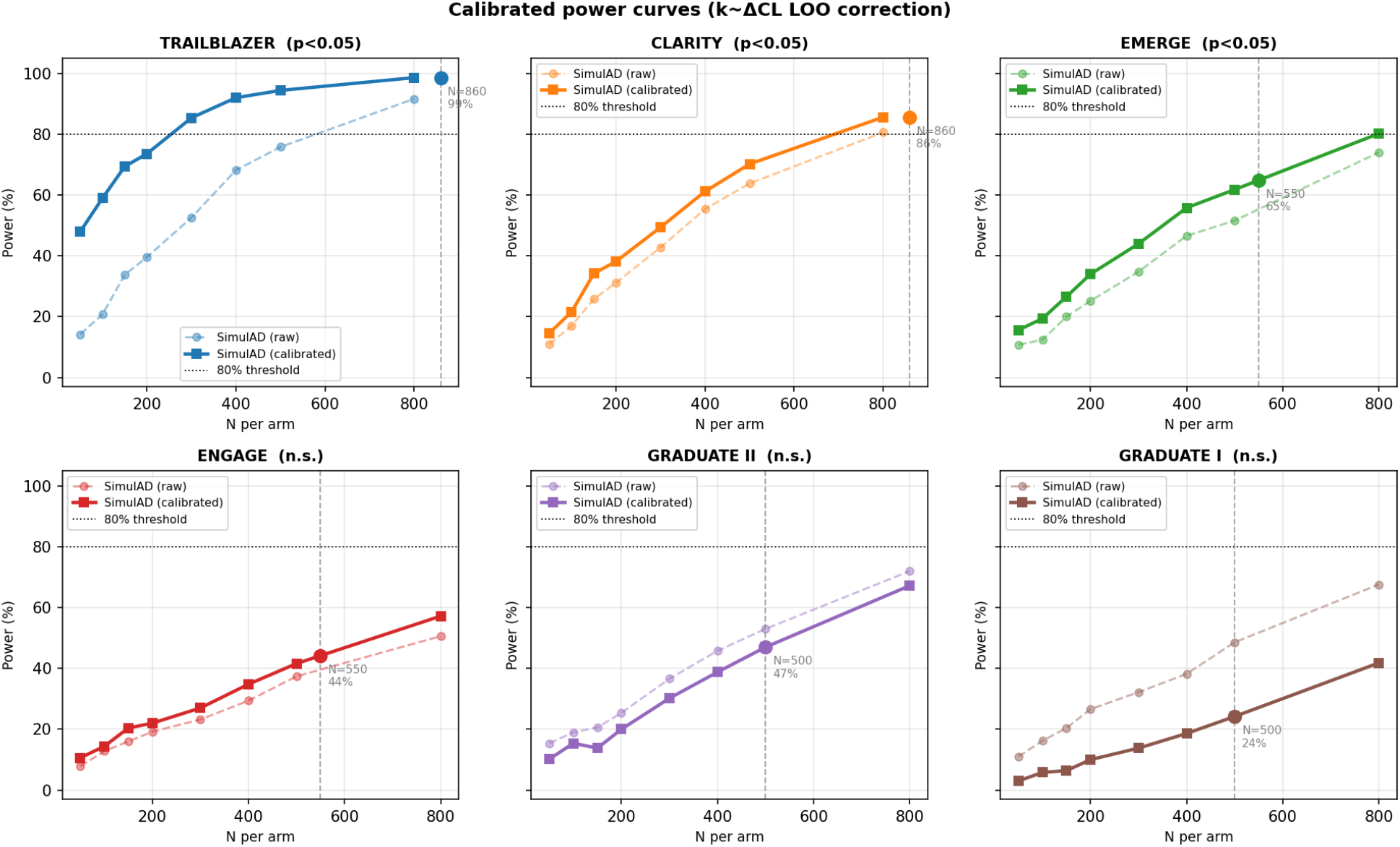
Uncalibrated power curves across six anti-amyloid trials. Empirical power (fraction of 500 repetitions with p < 0.05, LMM) as a function of N per arm, from raw SimulAD simulations without effect-size or SD calibration. Vertical ticks: actual enrolled N per arm.

**Table S5.** N_80_ and power at the enrolled N per arm for four calibration strategies. Oracle: N_80_ from published real effect and SD. Fully prospective: 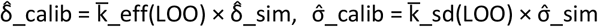. Semi-prospective: 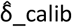 with published σ_real. Raw: uncalibrated 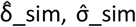. Power computed analytically (two-sample t-test, α = 0.05 two-sided). ENGAGE real effect ≈ 0 (n.s.); oracle N_80_ > 4000.

| Trial | $N_2$ enrolled | $N_{80}$ (per arm) | | | | Power at enrolled $N$ | | | |
| --- | --- | --- | --- | --- | --- | --- | --- | --- | --- |
|  |  | Oracle | Fully prosp. | Semi prosp. | Raw | Oracle | Fully prosp. | Semi-prosp. | Raw |
| TRAILBLAZER | 860 | 333 | 148 | 216 | 552 | 99% | 99% | 98% | 92% |
| CLARITY | 860 | 600 | 668 | 734 | 801 | 92% | 86% | 83% | 81% |
| EMERGE | 550 | 500 | 722 | 583 | 938 | 84% | 65% | 72% | 55% |
| ENGAGE | 550 | >4000 | 1349 | 1076 | 1525 | 5% | 44% | 52% | 40% |
| GRADUATE II | 500 | 3658 | 1324 | 1284 | 1093 | 18% | 47% | 48% | 53% |
| GRADUATE I | 500 | 1375 | 2269 | 2167 | 1259 | 39% | 24% | 26% | 49% |

### S6. K_eff – *Δ*CL calibration fit

Figure S6 shows the relationship between the per-trial effect-size calibration factor 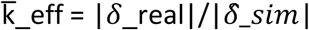 and the absolute amyloid reduction rate *Δ*CL/trial_duration for the five calibration trials (ENGAGE excluded as its real effect is near zero). A linear fit yields k = 0.5 + 0.48 × *Δ*CL/t (*R*^2^ = 0.85), indicating that trials with higher amyloid clearance rates require a proportionally larger correction to recover the real treatment effect from raw SimulAD estimates. The tight linear relationship supports the use of *Δ*CL/trial_duration as the predictor of 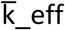, enabling fully prospective calibration from pre-existing amyloid clearance data.

**Figure S6a.**
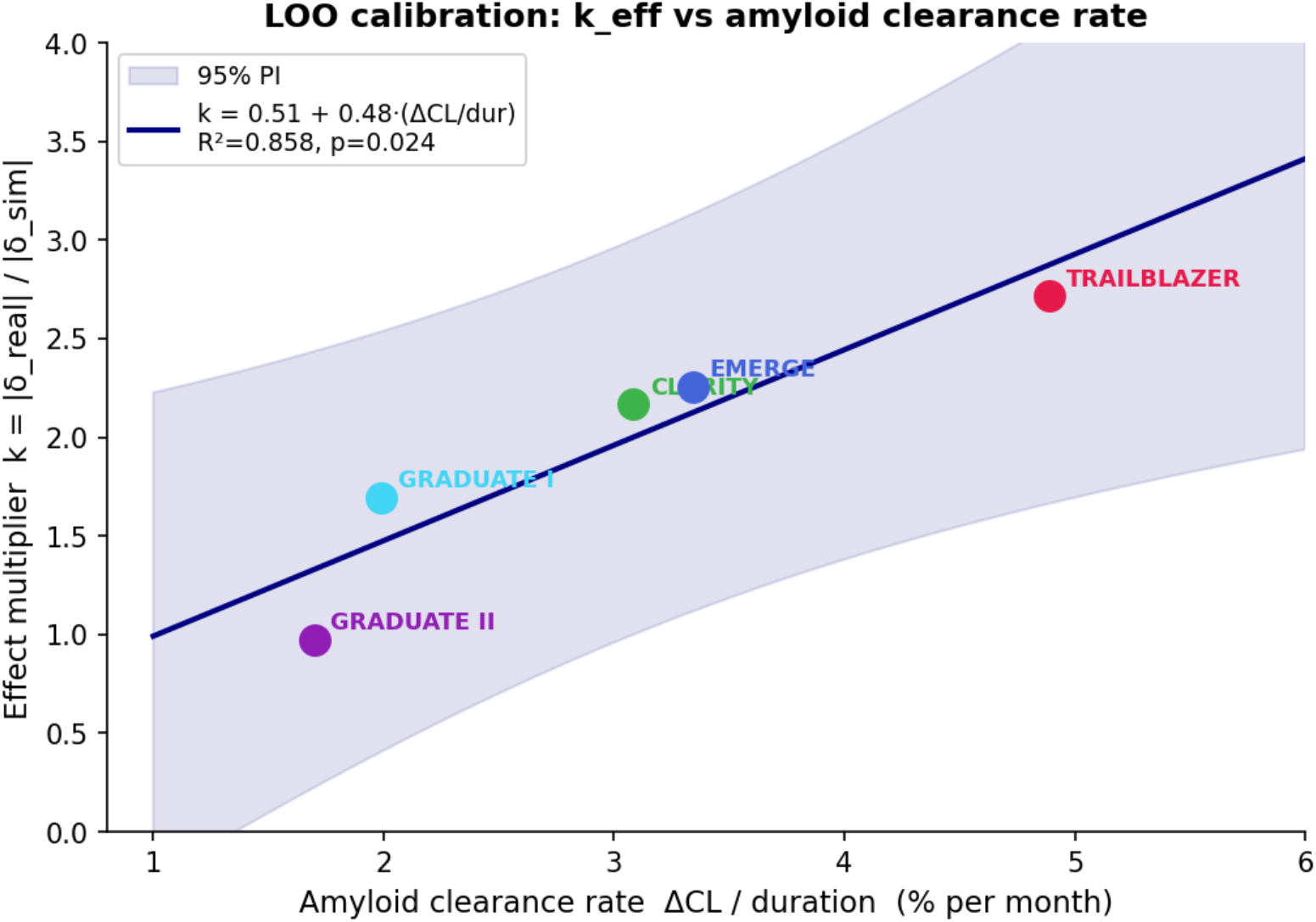
Effect-size calibration factor 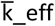 as a function of absolute amyloid reduction rate *Δ*CL/trial_duration. Filled circles: observed k per calibration trial. Solid line: full linear fit k = 0.5 + 0.48 × *Δ*CL/t (*R*^2^ = 0.85).

**Table S6a.** k∼ΔCL regression statistics for the calibration with and without TRAILBLAZER trial. Excluding TRAILBLAZER from the regression leads to minimal changes in slope and fit parameters, albeit with an increased prediction interval.

| Statistic | Full (5 trials) | No TRAILBLAZER (4 trials) |
| --- | --- | --- |
| Intercept | 0.5 | 0.05 |
| Slope | 0.48 | 0.68 |
| $R^2$ | 0.86 | 0.86 |
| Residual SE | 0.28 | 0.27 |
| Mean 95% PI width | 2.15 | 3.18 |

**Table S6b.** Fully prospective N_80_ using original LOO k_eff regression versus fully prospective N_80_ when TRAILBLAZER is excluded from all regressions.

| Trial | $\bar{k}_{\text{eff}}(\text{LOO})$ original | $\bar{k}_{\text{eff}}(\text{LOO})$ no TB | $N_{80}$ original | $N_{80}$ no TB |
| --- | --- | --- | --- | --- |
| TRAILBLAZER | 3.37 | 3.37 | 149 | 149 |
| CLARITY | 1.96 | 2.13 | 669 | 564 |
| EMERGE | 2.09 | 2.42 | 723 | 539 |
| GRADUATE II | 1.64 | 1.57 | 1325 | 1445 |
| GRADUATE I | 1.35 | 1.22 | 2270 | 2777 |

We note that TRAILBLAZER is an extreme point in the k∼ΔCL/trial_duration regression, reflecting its position at the extremity of the observed ΔCL range. Excluding TRAILBLAZER from the regression changes the slope by 41% (0.48 to 0.68 per ΔCL/t units, Supplementary Table S6a). The 95% prediction interval for k at a new trial’s ΔCL also widen (mean width ≈ 2.15 with all five trials; ≈ 3.2 without TRAILBLAZER), reflecting the intrinsic uncertainty of calibrating on five data points. Supplementary Table S6b shows however that the removal of TRAILBLAZER from the calibration set has negligible impact to the sample size estimation for all trials. The most notable changes are for EMERGE (-184 individuals) and GRADUATE I (+507 individuals), which are however still coherent with the statistical findings of the power analysis performed including TRAILBLAZER.

### S7. Multimodal cascade analysis: downstream biomarker effects of amyloid clearance

Table S7a reports the within-simulation subject-level correlations: for each individual subject we computed change scores ΔAV45, ΔTAU, and ΔVol at the primary endpoint, then computed Pearson r and Spearman ρ of ΔAV45 vs each downstream biomarker separately in the treated and placebo arms (N = 3,200 subjects per arm, 8 repetitions × 400 subjects).

**Table S7a.** Within-simulation subject-level correlations between individual amyloid change (ΔAV45 = AV45(BL) − AV45(endpoint)) and downstream biomarker changes (Δτ: entorhinal tau SUVR change; ΔVol: hippocampal volume change) at the primary endpoint, separately for treated and placebo arms. Positive Δ values denote reduction. Pearson r and Spearman ρ computed across N = 3,200 subjects per arm (8 repetitions × 400 subjects per trial). Pooled row: z-scored within each trial before pooling across the six trials (N = 19,200). All correlations are statistically significant (p < 0.001). The treatment-arm attenuation relative to placebo reflects treatment-driven amyloid variance partially decorrelated from natural-history biomarker trajectories.

| Trial | Treated arm |  |  |  | Placebo arm |  |  |  | N |
| --- | --- | --- | --- | --- | --- | --- | --- | --- | --- |
| | $\Delta\text{AV45 vs } \Delta\tau$ | | $\Delta\text{AV45 vs } \Delta\text{Vol}$ | | $\Delta\text{AV45 vs } \Delta\tau$ | | $\Delta\text{AV45 vs } \Delta\text{Vol}$ | | |
| | r | $\rho$ | r | $\rho$ | r | $\rho$ | r | $\rho$ | |
| TRAILBLAZER | +0.421 | +0.404 | -0.103 | -0.097 | +0.780 | +0.769 | -0.715 | -0.696 | 3200 |
| CLARITY | +0.395 | +0.376 | -0.164 | -0.155 | +0.780 | +0.767 | -0.714 | -0.694 | 3200 |
| EMERGE | +0.393 | +0.374 | -0.174 | -0.165 | +0.777 | +0.764 | -0.711 | -0.691 | 3200 |
| ENGAGE | +0.379 | +0.360 | -0.224 | -0.213 | +0.776 | +0.763 | -0.710 | -0.690 | 3200 |
| GRADUATE II | +0.591 | +0.577 | -0.276 | -0.265 | +0.575 | +0.558 | -0.675 | -0.660 | 3200 |
| GRADUATE I | +0.590 | +0.576 | -0.276 | -0.264 | +0.580 | +0.563 | -0.678 | -0.664 | 3200 |
| Pooled (z-scored) | +0.461 | +0.445 | -0.203 | -0.194 | +0.711 | +0.698 | -0.700 | -0.683 | 19200 |

All individual-trial correlations: $p < 0.001$ (\*\*\*). Pooled: z-scored within trial before pooling.

## Notes

### Competing Interest Statement

The authors have declared no competing interest.

### Funding Statement

This study was funded by the grant ANR-22-PESN-0016 (REWIND), and by the 3IA Cote d'Azur initiatives, including ANR-19-P3IA-0002 (3IA Cote d'Azur) and ANR-23-IACL-0001 (3IA Cote d'Azur 2030)

### Author Declarations

This study is based on the publicly available Alzheimer's Disease Neuroimaging Initiative (ADNI) data (https://adni.loni.usc.edu/). ADNI is funded by the National Institute on Aging (National Institutes of Health Grant U19 AG024904). The grantee organization is the Northern California Institute for Research and Education. ADNI was conducted in accordance with the Declaration of Helsinki and all applicable national regulations. All participants (or their legally authorized representatives) provided written informed consent prior to enrollment. The study protocol, any amendments, and all recruitment materials were reviewed and approved by the institutional review board (IRB) or ethics committee at each participating site. Any data sharing followed the approved data-use agreements and procedures to prevent re-identification. Safety monitoring was performed according to protocol; adverse events were reported to local IRBs and oversight bodies as required.

## Bibliography

1. Estimation of the global prevalence of dementia in 2019 and forecasted prevalence in 2050: an analysis for the Global Burden of Disease Study 2019. E, Nichols, et al. s.l. : The Lancet Public Health, 2022, The Lancet Public Health, pp. Feb 1;7(2):e105–25.

2. Why do trials for Alzheimer’s disease drugs keep failing? A discontinued drug perspective for 2010-2015. D, Mehta, et al. 6, 2017, Expert opinion on investigational drugs, Vol. 26, pp. 735–9.

3. Lecanemab in early Alzheimer’s disease. CH, Van Dyck, et al. 1, 2023, New England Journal of Medicine, Vol. 388, pp. 9–21.

4. Donanemab in early symptomatic Alzheimer disease: the TRAILBLAZER-ALZ 2 randomized clinical trial. JR, Sims, et al. 6, 2023, Jama, Vol. 330, pp. 512–27.

5. Two randomized phase 3 studies of aducanumab in early Alzheimer’s disease. SB, Haeberlein, et al. 2, 2022, The journal of prevention of Alzheimer’s disease, Vol. 9, pp. 197–210.

6. NIA-AA research framework: toward a biological definition of Alzheimer’s disease. CR, Jack Jr, et al. 4, 2018, Alzheimer’s & dementia, Vol. 14, pp. 535–62.

7. The amyloid hypothesis of Alzheimer’s disease: progress and problems on the road to therapeutics., Vol.J, Hardy and DJ, Selkoe. 5580, 2002, Science, Vol. 297, pp. 353–6.

8. Hypothetical model of dynamic biomarkers of the Alzheimer’s pathological cascade., Vol.CR, Jack, et al. 1, 2010, The Lancet Neurology, Vol. 9, pp. 119–28.

9. Clinical trial simulation: a review., Vol.N, Holford, SC, Ma and BA, Ploeger. 2, 2010, Clinical Pharmacology & Therapeutics, Vol. 88, pp. 166–82.

10. Disease progression meta-analysis model in Alzheimer’s disease., Vol.K, Ito, et al. 1, 2010, Alzheimer’s & Dementia, Vol. 6, pp. 39–53.

11. An event-based model for disease progression and its application in familial Alzheimer’s disease and Huntington’s disease. HM, Fonteijn, et al. 3, 2012, NeuroImage, Vol. 60, pp. 1880–9.

12. Estimating long-term multivariate progression from short-term data. MC, Donohue, et al. S400, 2014, Alzheimer’s & Dementia., Vol. 10, pp. 10.

13. Uncovering the heterogeneity and temporal complexity of neurodegenerative diseases with Subtype and Stage Inference. AL, Young, et al. 1, 2018, Nature communications, Vol. 9, pp. 4273.

14. A Bayesian mixed-effects model to learn trajectories of changes from repeated manifold-valued observations. JB, Schiratti, et al. 133, 2017, Journal of Machine Learning Research, Vol. 18, pp. 1–33.

15. Simulating the outcome of amyloid treatments in Alzheimer’s disease from imaging and clinical data. Brain communications. C, Abi Nader, et al. 2, 2021, Brain communications, Vol. 3, p. fcab091.

16. SimulAD: A dynamical model for personalized simulation and disease staging in Alzheimer’s disease. C, Abi Nader, et al. 2022, Neurobiology of Aging, Vol. 113, pp. 73–83.

17. Two phase 3 trials of gantenerumab in early Alzheimer’s disease. RJ, Bateman, et al. 20, s.l. : New England Journal of Medicine., 2023, Vol. 389. 1862–76.

18. Neuro-Dynamic Quantitative Systems Pharmacology (Qsp) Model Supports Continued Lecanemab Treatment With Maintenance Dosing For Alzheimer’s Disease. Y, Cao, et al. s.l. : Alzheimer’s & Dementia, 2024. p.e092093.

19. The Alzheimer’s disease neuroimaging initiative. SG, Mueller, et al. 4, s.l. : Neuroimaging Clinics, 2005, Vol. 15. 869–77.

20. Auto-Encoding Variational Bayes. Kingma, D.P. and Welling, M. 2014 : 2nd International Conference on Learning Representations. ICLR.

21. Neural ordinary differential equations. Chen RT, Rubanova Y, Bettencourt J, Duvenaud DK. s.l. : Advances in neural information processing systems, 2018. 31.

22. Adam: A Method for Stochastic Optimization. Kingma, DP and Ba, J. s.l. : International Conference on Learning Representations (ICLR), 2015.

23. Statsmodel. [Online] [Cited: March 28, 2026.] https://www.statsmodels.org/stable/index.html.

24. Scikit-learn: Machine learning in Python. Pedregosa, F., et al. 282, s.l. : Journal of machine Learning research, 2011, Vol. 12.

